# Assessment of effective isolation, safe burial and vaccination optimal controls for an Ebola epidemic model

**DOI:** 10.1101/2020.06.03.20121319

**Authors:** Arsène Jaurès Ouemba Tassé, Berge Tsanou, Jean M.-S. Lubuma, Jean Louis Woukeng

## Abstract

Since 1976 many outbreaks of Ebola virus disease have occurred in Africa, and up to now, no treatment is available. Thus, to fight against this illness, several control strategies have been adopted. Among these measures, isolation, safe burial and vaccination occupy a prominent place. In this paper therefore, we present a model which takes into account these three control strategies as well as the difference in immunological responses of infected by distinguishing individuals with moderate and severe symptoms. The sensitivity analysis of the control reproduction number suggests that, the vaccination response works better than the two other control measures at the beginning of the disease, and there is no need to vaccinate people in order to overcome Ebola. The global asymptotic dynamic of the model with controls is completely achieved exhibiting a sharp threshold behavior. Consequently, the global stability of the endemic equilibrium when the control reproduction number is greater than one, calls for the implementation of the three responses optimally. Thus, we define, analyze and implement different optimal control problems which minimize the cost of controls and the number of infected individuals through vaccination, isolation and safe burial. The obtained results highlight that, in order to mitigate Ebola outbreaks, an optimal vaccination strategy has more impact than the combination of isolation and safe burial responses. Therefore, the broad recommendation is that: if for any disease outbreak, the vaccine can be available quickly, one should think of implementing large vaccination programs rather than any other control measures.

## 1 Introduction

In December 2013 and August 2018, two deadliest of Ebola virus disease (EVD) outbreaks began in Guinea and DRC (Democratic Republic of Congo) respectively. About 11 000 deaths have been registered during the first one, and at this time the second has caused more than 2200 deaths [74,22]. EVD symptoms generally include fever, severe headache, muscle aches, weakness, vomiting, diarrhea, stomach pains, loss of appetite and at times hemorrhagic bleeding, which can lead to death [1,59]. It is documented that, some EVD infected individuals may present moderate symptoms, while others severe symptoms [20, 51, 55, 61]. In fact, the difference in immunological responses may be related to host genetics and/or partial immunity due to previous infection with a related virus [20, 51, 55, 31]. EVD can be transmitted through close contacts with susceptible individuals and bodily fluids such as blood, stools, vomiting, saliva, urine, sperm, maternal milk,… of infected ill or dead persons [1, 16, 59].

The Ebola-deceased individuals can transmit the disease during mourning or traditional funerals which include rites and others practices on their corpses [8]. Thus, to impede the transmission by the deceased, education of people to safe burial has been at the center of all battles against EVD. However, due to the attachment to cultural norms, many people have contracted EVD through manipulation of Ebola-deceased bodies [48]. Moreover, the lack of adequate equipment has caused the infection of many health workers in the Ebola treatment units (ETU) and hospitals. Thus, responding to the EVD spreading by isolating people has not been always perfect [44,73]. Furthermore, up to now, neither treatment nor vaccine is ratified by the World Health Organization (WHO). However, an experimental vaccine named rVSV-ZEBOV developed in the course of 2013-2015 outbreak has demonstrated a great efficacy [5,47], and is used to control the 2018-2020 EVD outbreak in DRC through ring vaccination [15,79,80]. Precisely, at the date of Mai 2019 more than 223 000 people have already received the vaccine during the ongoing Ebola outbreak, and since then this number has not evolved significantly up to date [77]. Therefore, the public health officials and the scientific community should considered the combination of vaccination and former control strategies such as isolation and safe burial in the fight against EVD [15]. Such a battle gives floor to a multi-disciplinary research activity among which, mathematical modeling and control theory play.

Essentially, mathematical models may help the medical community and decision makers to evaluate the effectiveness of different approaches for bringing an epidemic under control [34]. In this regard, since 2014 many models for EVD were focused on control strategies such as contact tracing, isolation, safe burial and vaccination. For instance, in [12], the authors proposed a model which includes vaccination and heterogeneity of the risk of becoming infected with EVD. Contrarily to [12], in [11], the authors proposed an EVD model with isolation, where the Ebola-deceased individuals are divided in two compartments: those who can transmit the disease and those who can not transmit it, since they are safely buried. They computed the basic reproduction number and evaluated the efficiency of different control strategies to reduce its value. But, it seems confusing to separate the compartment of hospitalized individuals from that of isolated, since infected people in both classes are settled in a treatment center (which are not opened to the general public [58]). Furthermore, in [23], the authors investigated the usefulness of quarantine and isolation to mitigate EVD outbreaks. They showed numerically that, a good implementation of quarantine and the reduction of contacts between quarantined and infectious individuals can extensively reduce the level of the disease and may eliminate it. However, in their work, the authors assumed that the recovery rate of infected individuals is the same whether they are in hospitals or not. This may be not always the case, and we believe that, this assumption should relaxed. Beside, in [62, 66], the authors proposed models with vaccination and isolation, where in the first paper, isolated individuals can transmit the disease but as compared to infected non-isolated individuals, they computed the basic reproduction number and performed the sensitivity analysis of their model. This analysis shows that, the efficacy of isolation and the time from death to burial have the most influence on the basic reproduction number than all others parameters. While, in the second paper, they assessed the efficiency of vaccination and quarantine in the fight against EVD. Our concern in [62] is that fact that, the authors assumed that, the latent and symptomatic individuals are isolated in the same compartment, since EVD is unlikely be transmitted during the incubation period. In the other hand, in [66], the authors did not take into account the infectivity potential of Ebola-deceased individuals. These later drawbacks can also be highlighted in the models with isolation proposed in [19, 36, 43, 53, 83]. In [51], the authors proposed an interesting model with isolation which takes into account the transmissibility of individuals with moderate symptoms, severe symptoms and Ebola-deceased. However, they did not consider the fact that, some infected may progress from moderate symptoms to severe symptoms. Moreover, it is not clearly explained or motivated why those individuals who show moderate symptoms can not recover.

In this paper, we focus on the above mentioned three main prevention strategies which are: vaccination, imperfect isolation and safe burial by combining them in a single model, which additionally fills many (if not all) the drawbacks highlighted the above-mentioned papers. However, one should not lose sight that, contact tracing is another efficient control measure for Ebola which has already been investigated by us and other researchers [9, 29, 45]. Our propounded model is qualitatively and quantitatively analyzed. We prove the existence of a vaccination threshold which can determine whether the disease will dies out or be endemic. Through sensitivity analysis of the control reproduction number, we show that, vaccination is the most influential response to mitigate EVD at the beginning of the disease outbreak; suggesting that, for long lasting EVD epidemics, vaccinating people might not be the panacea and urges additional control measures. On the other hand, we successfully implement optimal control problems that aim to minimizing the number of infected and Ebola-deceased individuals through vaccination alone and isolation-safe burial altogether.

The rest of this paper is organized as follows. In Section 2, the model is built in a comprehensive manner. The resulted system of differential equations is derived and the preliminary results are given. In Section 3, the sensitivity analysis is addressed. The global asymptotic behavior of the model is provided through the global stability analysis of equilibria in Section 4. Optimal control problems are analyzed and numerically implemented and commented in Section 5, and the conclusion and perspectives are provided in Section 6.

## 2 Model formulation and mathematical preliminary results

### 2.1 Main assumptions

1. The isolation is not always effective.
2. Some EVD infected individuals who died outside a medical center/Ebola treatment units (ETUs) are safely buried when their family members are educated accordingly.
3. The asymptomatic infected individuals do not transmit EVD (even though there is evidence of asymptomatic carriers [30], the very low level of virus detected in them do not indicate a significant source of transmission [40, 42]).
4. Ring vaccination is considered throughout this paper, so that susceptible individuals are at high risk of infection [78].
5. Hospitals and ETUs are isolation settings, and severely infected individuals can only recover at hospitals/ETUs.
6. We assume that all the Ebola infected individuals who succumb at the hospitals/ETUs are safely buried by well-trained personal.

### 2.2 Model variables

Our population is subdivided into the following mutually exclusive classes:

- *S*(*t*): Individuals at high risk of becoming infected. We assume that this class incorporates susceptible health workers (at the hospitals or in the Ebola treatment units), and uninfected visitors at hospitals/ETUs who take care for their sick ones.
- *J*(*t*): Infected individuals with moderate symptoms outside of the hospitals/ETUs. We recall that those symptoms include fever, sore throat, muscular pain and headaches [26].
- *I*(*t*): Infected individuals with severe symptoms (i.e vomiting, bloody diarrhea and rash) outside the hospitals/ETUs [1, 26, 59].
- *H*(*t*): Infected individuals who are under treatment (in hospitals/ETUs). We called them interchangeably isolated individuals.
- *D*(*t*): Ebola-deceased individuals. This class is an infectious class which gathers all the Ebola infected individuals who die anywhere except at the hospitals/ETUs, either naturally (or due any other comorbidity) or due EVD. The rationale of considering this class as such is threefold: (1) It is well documented that Ebola-deceased individuals are infectious if not properly handled. (2) Once an individual is infected with EVD and dies, its death is usually attributed to EVD, whether or not he/she dies naturally or due to EVD (actually, it is very difficult to determine the type of death in this case). (3) Ebola-deceased individuals at the hospitals/ETUs are properly handled by well-trained personal and therefore can no longer transmit the disease.
- *R*(*t*): Recovered individuals. Even though up to now, there is no available treatment for EVD, some patients have recovered after receiving supportive treatments [3]. Moreover, since it is documented that recovered individuals develop antibodies that last for at least 10 years [9, 35], we assume that they are permanently immune.

Let denote the overall total population (including dead individuals) and the alive total population, respectively by:

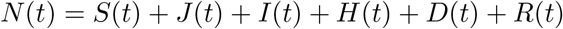

and

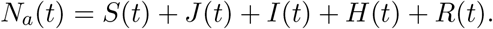

#### Remark 2.1

*The fact that the individuals of class J exhibit moderate symptoms do not formally means that their effective transmission rate is lower than that of individuals on compartment I. Let’s recall that, the effective transmission rate is the product of the contact rate and the. probability of being infected during that contact. Therefore, even though this probability can be high for the individuals of the I-compartment, the contact rate between susceptible and individuals of that class can be lower as compared to that in class J. Moreover, the individuals of compartment I can have an hospitalization rate greater than that of compartment J. Thus, individuals in J might, greatly contribute to the transmission of Ebola, and should not be neglected as far as EVD control is concerned*.

### 2.3 Derivation of model equations

Susceptible individuals in *S* are recruited at a constant rate *π* by immigration/births, and vaccinated at rate *υ*. Among vaccinated individuals, a proportion *θ* acquires immunity (since no vaccine is 100% effective [76]). The other susceptible individuals can be infected (i)-through contact with individuals in *J* and *I* classes at rates *β* and *βν*_1_ respectively, (ii)-through contact with (1 − *σ*_1_)H individuals who are not effectively isolated (where *σ*_1_ represents the proportion of infected who are effectively isolated) at rate *εβ*, or (iii) through contact with (1 − *σ*_2_)*D* Ebola-deceased individuals who failed safe burial (where *σ*_2_ is the proportion of Ebola-deceased individuals safely buried) at rate *βν*_2_. Since human immune system reacts differently, a proportion *p* of infected individuals may present moderate symptoms while the remaining proportion exhibits severe symptoms. The individuals in class *J* can recover (without being in at hospital/ETU) at rate *γ_j_*; die due to the disease at rate *δ_j_*; go to the hospital at rate *η_j_* or progress to compartment *I* at rate *α*. Those in compartment *I* can pass away at rate *δ*_i_ or become hospitalized at rate *η_i_* Hospitalized individuals recover at rate *γ_h_* or die at rate *δ_h_*. The alive population is affected at all stages by the common natural mortality rate

The model parameters and their biological meanings are summarized in Table 1, the transmission diagram of the disease is displayed in Figure 1, and the resulted system of nonlinear ordinary differential equations is as follows:

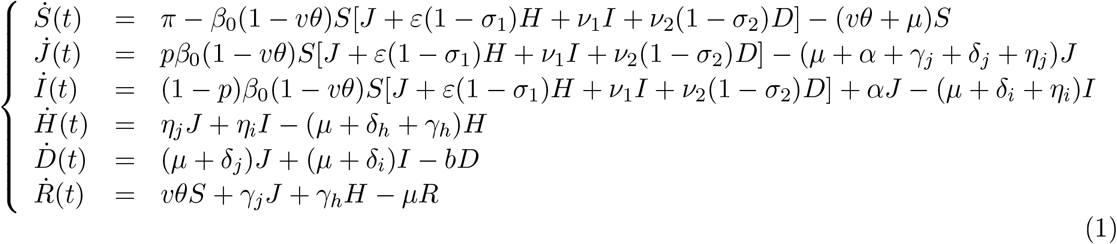

In order to simplify the notations and avoid lengthy expressions, we define the parameters:

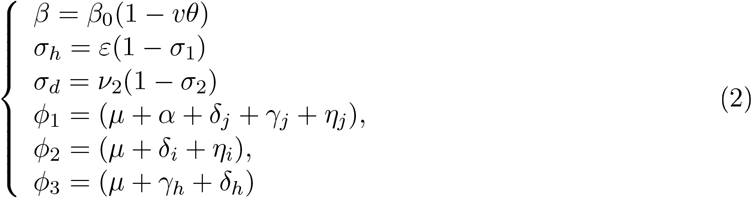

Thus, model (1) becomes

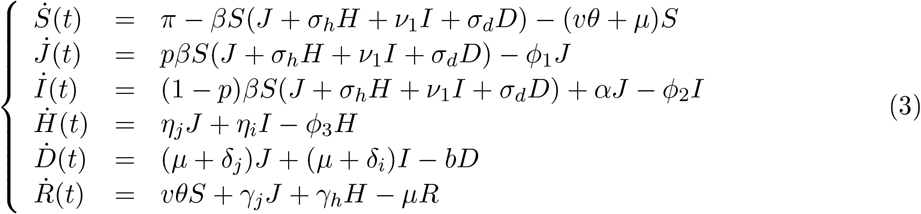

The mathematical proofs of the main theoretical results are provided in the Appendix Section and the interested reader is refereed to it for details.

**Figure 1:**
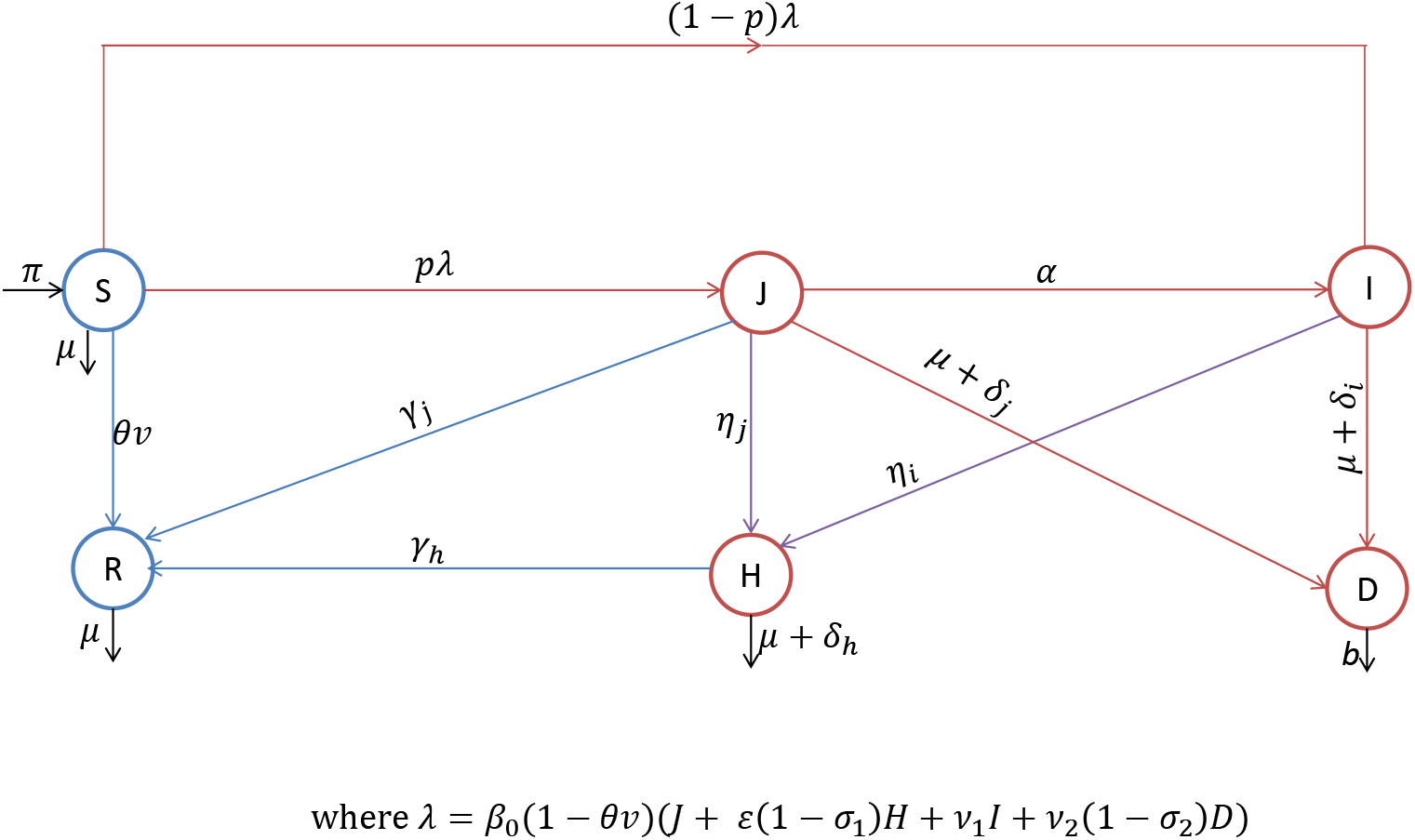
Transmission diagram of the model

### 2.4 Well-posedness of model (3)

#### Theorem 2.2

*The positive orthant* 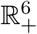 *is positively invariant under the flow of* (1). *Precisely, if S*(0) > 0, *J*(0) ≥ 0, *I*(0) ≥ 0, *H*(0) ≥ 0, *D*(0) ≥ 0, *R*(0) ≥*0 then* ∀ *t* ≥ 0, S(t) *>* 0, *J*(*t*) ≥ 0, *I*(*t*) ≥ 0, *H*(*t*) ≥ 0, *D*(*t*) ≥ 0, *R*(*t*) ≥ 0.

The proof of this theorem is given to Appendix A. Moreover, using the Gronwall lemma, one can easily prove the following result.

#### Theorem 2.3

*Let’s suppose the initial conditions of system* (1) *are as in Theorem 2.2*.

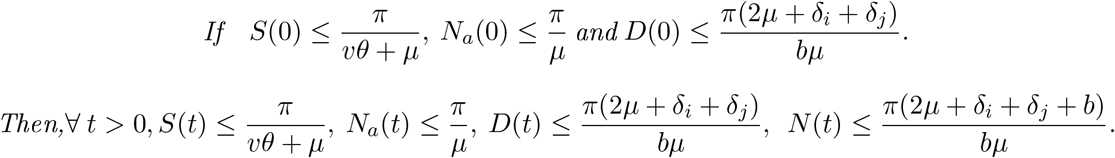

A direct consequence of Theorem 2.2 and Theorem 2.3 is the following result, which ensures the existence and uniqueness of global (in time) solution of system (1).

#### Theorem 2.4

*The model (*1*) is a dynamical system on*

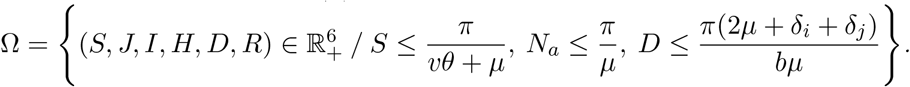

**Table 1:**
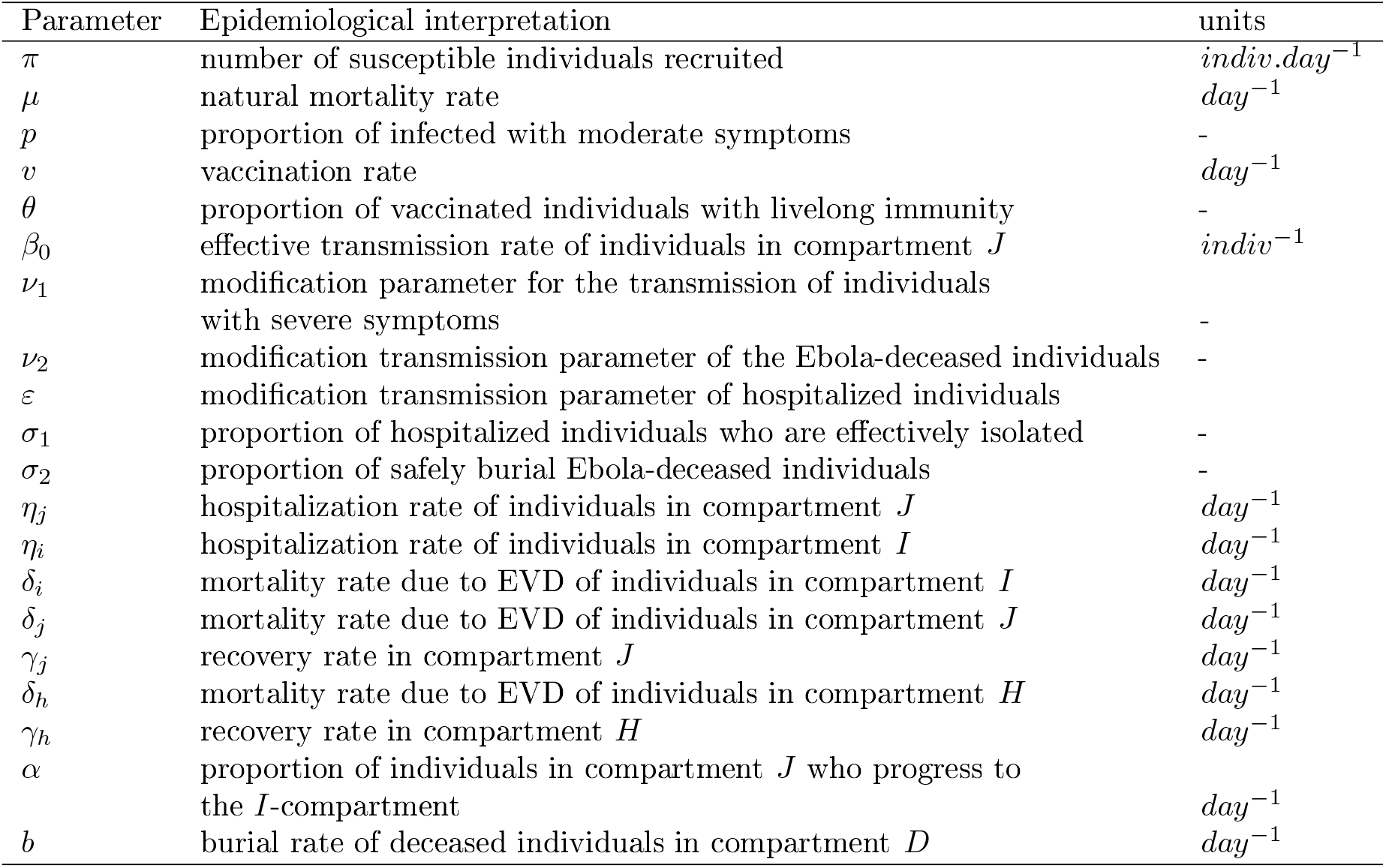
Parameters and epidemiological interpretation of model (1).

| Parameter | Epidemiological interpretation | units |
| --- | --- | --- |
| $\pi$ | number of susceptible individuals recruited | $indiv.day^{-1}$ |
| $\mu$ | natural mortality rate | $day^{-1}$ |
| $p$ | proportion of infected with moderate symptoms | - |
| $v$ | vaccination rate | $day^{-1}$ |
| $\theta$ | proportion of vaccinated individuals with lifelong immunity | - |
| $\beta_0$ | effective transmission rate of individuals in compartment $J$ | $indiv^{-1}$ |
| $\nu_1$ | modification parameter for the transmission of individuals with severe symptoms | - |
| $\nu_2$ | modification transmission parameter of the Ebola-deceased individuals | - |
| $\varepsilon$ | modification transmission parameter of hospitalized individuals | - |
| $\sigma_1$ | proportion of hospitalized individuals who are effectively isolated | - |
| $\sigma_2$ | proportion of safely burial Ebola-deceased individuals | - |
| $\eta_j$ | hospitalization rate of individuals in compartment $J$ | $day^{-1}$ |
| $\eta_i$ | hospitalization rate of individuals in compartment $I$ | $day^{-1}$ |
| $\delta_i$ | mortality rate due to EVD of individuals in compartment $I$ | $day^{-1}$ |
| $\delta_j$ | mortality rate due to EVD of individuals in compartment $J$ | $day^{-1}$ |
| $\gamma_j$ | recovery rate in compartment $J$ | $day^{-1}$ |
| $\delta_h$ | mortality rate due to EVD of individuals in compartment $H$ | $day^{-1}$ |
| $\gamma_h$ | recovery rate in compartment $H$ | $day^{-1}$ |
| $\alpha$ | proportion of individuals in compartment $J$ who progress to the $I$-compartment | $day^{-1}$ |
| $b$ | burial rate of deceased individuals in compartment $D$ | $day^{-1}$ |

### 2.5 Equilibria of model (3) and control reproduction number

For clarity, we set:

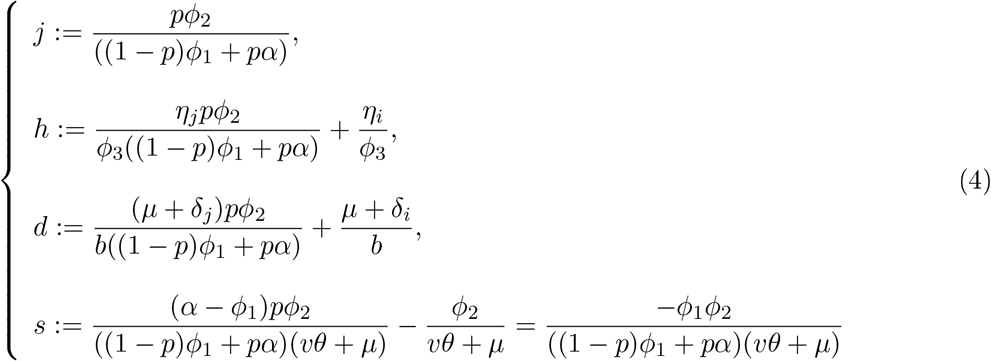

Using the method described in [70], one can easily show that, the control reproduction number 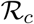 of model (3) reads:

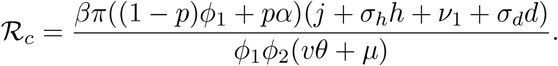

#### Remark 2.5

*Note that*, 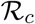*, can be rewritten in the form of four contributions as follows:*

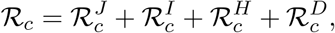

*where*

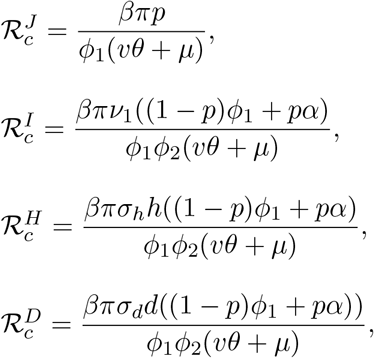

*are respectively the contributions of infected, individuals in J, I, H and D classes, to the control reproduction number. This decomposition of* 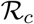 *suggests that, a good isolation and education of people to practice safe burial can extensively reduce the value of* 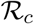 *(sinee σ_d_ and σ_h_ will be reduced). In the particular case, where, isolation is effective, and all Ebola-deceased individuals are safely buried (i.e σ_i_* = *σ_2_* = 1), 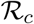 *becomes*

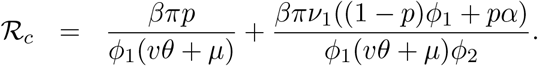

Moreover, we have the following result whose proof is postponed to Appendix B.

#### Theorem 2.6

*If the control reproduction number* 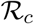 *is less than* 1*, model* (1) *has a unique equilibrium: the disease-free equilibrium* 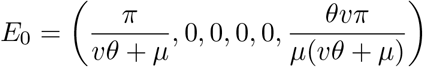

*Conversely, if* 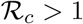*, model* (1) *has two equilibria: the DFE and a unique endemic equilibrium E*_1_ = (*S*^*^, *J*_*_,*I*^*^,*H^*^,D^*^,R^*^) which components are given by:*

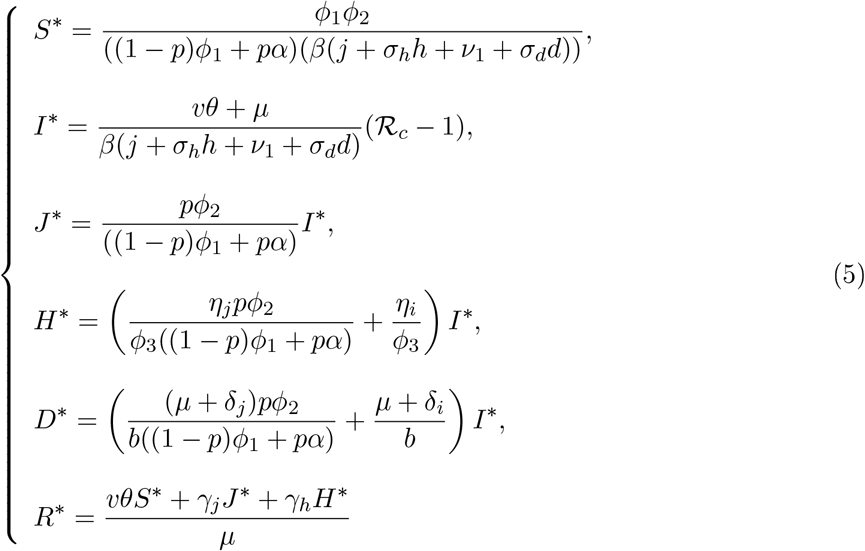

Thanks to [70], the following result is straightforward.

#### Lemma 2.7

*If* 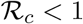*, the DFE is locally asymptotically stable. If* 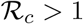*, the DFE is unstable*.

The epidemiological interpretation of Lemma 2.7 is that, EVD can be eliminated in the community when 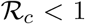 if the initial sizes of the different sub-populations of model (3) are in the basin of attraction of the DFE *E_0_*. But for a better control of the disease, the global asymptotic stability of the DFE need to be established. This will be done shortly. In the meantime, let’s recall that, the control reproduction number 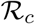 is the average number of secondary infections caused by a single infectious individual in an entirely susceptible population during its entire infectious period despite control measures [9]. Lemma shows that, 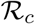 is a threshold which can determine if the disease will be spread or not. Thus, reducing its value, is a means to mitigate or even eliminate the EVD. It can be therefore important to determine among model parameters those who mostly influence its value.

## 3 Sensitivity analysis of the control reproduction number

### 3.1 Sensitivity analysis of the control reproduction number

The sensitivity analysis is important in designing control strategies that can be chosen at the onset of EVD outbreak, since it may help us to identify among parameters the most influential. To see how a small perturbation made to a parameter *q* affects the control reproduction number 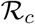, we define the sensitivity index of 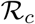 for *q* as follows [9, 13, 24, 38]:

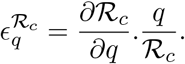

The sensitivity indices of 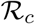 at the baseline values of model (1) are displayed in Table.

It is well known that, 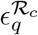 is positive if 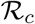 is increasing with respect to *q* and negative if 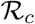 is decreasing with respect to *q* [9]. Thus, Table, suggests that *β_0_,ν_2_*, and *σ*_1_ are the most influential parameters which contribute to increase 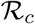; while *θ,υ,σ*_1_*,σ_2_* contribute to its decreasing. Thus, at this point and at the onset of EVD, vaccination seems to be the best control strategy among those which are to be addressed optimally in this model. However, in order to fasten the elimination of EVD, it is important to investigate the relevance its combination with the two other control measures mentioned earlier.

### 3.2 Efficiency of the combination of control strategies at the onset of the outbreak

Our model addresses three control strategies: the vaccination, isolation and safe burial, and the impact of each of them on the control reproduction number was presented in the previous subsection. Here, we focus on the combination of two of them. In this vein, we construct in Figure 2 the contour plot of 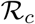 when (i) vaccination is combined with isolation; (ii) vaccination is combined with safe burial; and (iii) safe burial is combined with vaccination. Here, the parameters values used are given by Table 2. Figure 2 shows that, vaccination alone can not overcome EVD. Its combination with isolation or safe burial might serves the purpose. Precisely, when the values of *σ*_1_ or *σ*_2_ is greater than 0.6, EVD will be eliminated. This suggests that, EVD can be easily controlled in the cities/countries where people do not practice harmful rites on corpses, or where hospitals/ETUs are equipped with protective materials for health workers. However, in other to predict the long run dynamics of model 3 that could guide and justify the control strategies, it is advisable to know precisely some model features from the theoretical aspect.

**Table 2:** Sensitivity indices and parameters values for simulations.

| Parameter | Baseline value | Source | Sensitivity index |
| --- | --- | --- | --- |
| $\beta_0$ | 0.003 | [8] | 1 |
| $\nu_2$ | 0.0012/0.003 | [8] | 0.31 |
| $\nu_1$ | 0.2 | Assumed | 0.26 |
| $\varepsilon$ | 0.3 | Assumed | 0.27 |
| $p$ | 0.03 | Assumed | 0.127 |
| $\pi$ | 100 | [6] | 1 |
| $\delta_i$ | 0.5 | [8] | -0.263 |
| $\delta_j$ | 0.025 | Assumed | -0.028 |
| $\delta_h$ | 1/10.07 | [36] | -0.155 |
| $\gamma_h$ | 0.06 | [8] | -0.09 |
| $\gamma_j$ | 0.0314862 | [45] | -0.022 |
| $\sigma_2$ | 0.523 | [9] | -0.27 |
| $\sigma_1$ | 0.523 | Assumed | -0.293 |
| $b$ | 1/2.01 | [2, 5, 56] | -0.247 |
| $v$ | 0.4 | [12] | -1.13 |
| $\theta$ | 0.54 | [62] | -2.07 |
| $\mu$ | 10.17/ 1000 | [9] | -0.028 |
| $\eta_i$ | 0.25 | [35] | 0.07 |
| $\eta_j$ | 0.125 | Assumed | -0.119 |
| $\alpha$ | 0.025 | Assumed | -0.07 |

## 4 Global stability of equilibria

Let’s denote

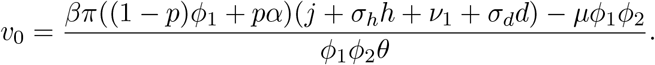

### Theorem 4.1

*If υ* ≥ *υ_0_ then the DFE of model* (3) *is globally asymptotically stable*.

The proof of this theorems is provided in Appendix C.

The biological interpretation of Theorem 4.1 is that, if one can vaccinate people at a rate *υ* greater than *υ*_0_, then EVD will be eliminated. In particular, if it happens that *υ*_0_ is negative, then no need to vaccinate because, any rate of vaccination will be sufficient to eliminate the disease.

On the other hand, following Table 2, the control reproduction number is not very sensitive to the parameter *α*. Therefore, the influence of this transfer rate from individuals with moderate symptoms to those with severe symptoms can be neglected. This was probably the reason (even though they did not highlight it) why the authors of the model proposed in [51] did not take into consideration this transfer rate. Moreover, we have shown that under the latter simplification (*α* = 0), the disease persists globally in the population, as highlighted in the following result.

### Theorem 4.2

*Set*

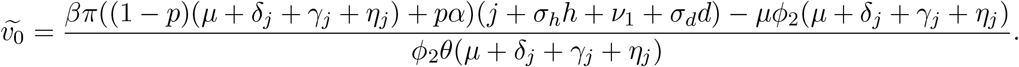

*If* 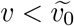, *then the endemic equilibrium of model* (3) *is globally asymptotically stable*.

**Figure 2:**
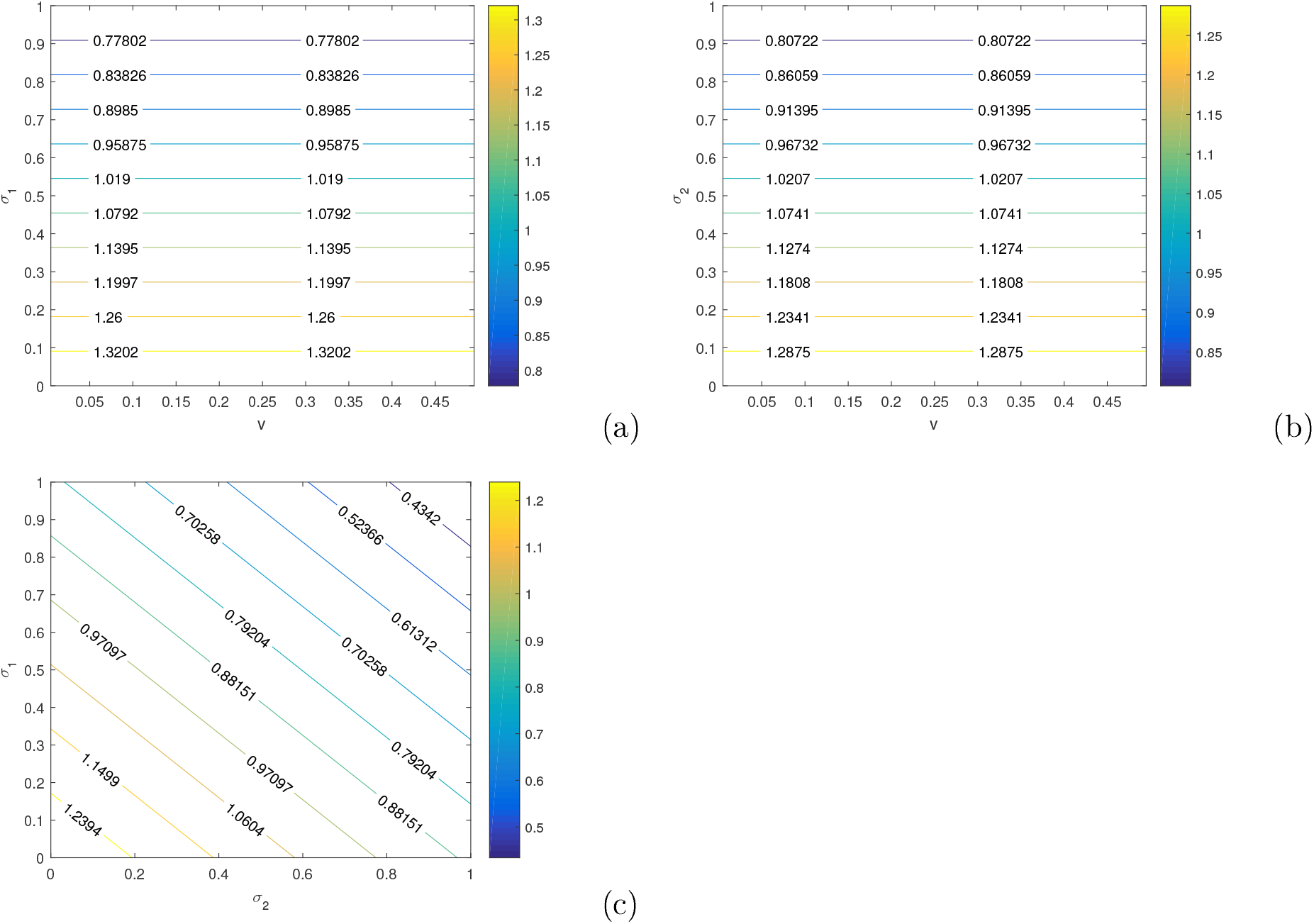
Contour plots of the control reproduction number in the, (*σ*_1_,*υ*), (*σ*_2_,*υ*) and (*σ*_1_, *σ*_2_) espaces, respectively when *π* = 70, The other parameters are as in Table 2.

The proof of this theorem is established in Appendix C.

The global persistence of EVD established in Theorem 4.2, whenever the control reproduction number is greater than one urges the reinforcement of control measures, which are going be addressed in the next section by setting and implementing targeted optimal control problems.

## 5 Optimal control

Since 1976, isolation and safe burial have been the main responses to EVD spreading. However, during the 2018-2019 RDC outbreak, ring vaccination was experimented [15, 79, 80]. It can be therefore important to explore the optimal impact of this novel EVD control measure on the dynamics of EVD and to compare its impact with that of the former control measures (isolation, safe burial).

### 5.1 Optimal vaccination

The main purpose of vaccination is to mitigate the propagation of a disease through the reduction of the number of susceptible individuals in the population. We assume that vaccination takes place at an unknown time dependent rate *υ*(*t*). Thus, model (1) becomes:

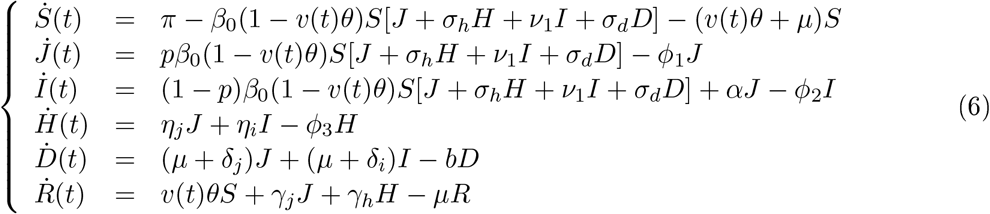

According to model (6), we remark that, the minimization of infected individuals in *J* and *I* classes leads to a reduction of hospitalized and Ebola-deceased individuals. Thus, to find an optimal vaccination strategy that minimizes the whole infectious population, we define the simple cost functional

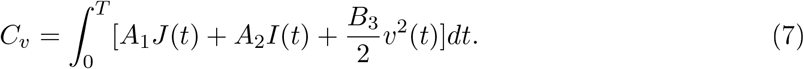

In Equation (7), *T* is the maximum duration of the vaccination process, *A*_1_*, A*_2_ are the weight factors associated to infected individuals with moderate symptoms and with severe symptoms, respectively; while *B*_3_ is the cost attached to the vacination process. The function *υ*(*t*) is assumed to be Lebesgue measurable.

Set

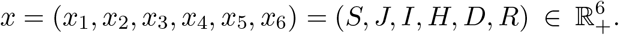

Our optimal control problem here consists to find the optimal trajectory *x^*^* associated to the optimal control *υ^*^* satisfying the control system (6) that minimizes the objective functional (7) over the set

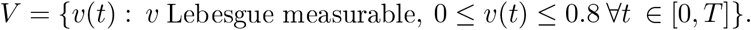

#### Theorem 5.1

*There exists υ^*^* ∈ *V so that the objective functional in* (7) *is minimized. Moreover, for small enough T, υ^*^ is unique*.

##### Proof

The existence of an optimal control *υ*^*^ and the associated optimal trajectories *x*^*^ is guaranteed by the convexity of the integrand of the cost function with respect to the control *υ* [71]. Besides, the Lipschitz property of the state system with respect to state variables and the boundedness of the state solutions of system (1) ensure the uniqueness of the control for sufficiently small *T* [38].

Moreover, following the Pontryagin’s maximum principle, there exists a function [5, 14, 41]

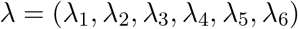

called the adjoint vector satisfying the system

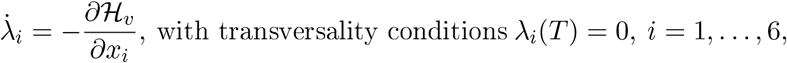

where 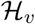 is the Hamiltonian defined bv:

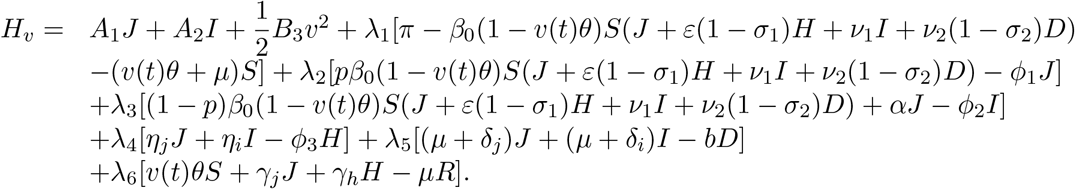

The adjoint variables satisfy the system below:

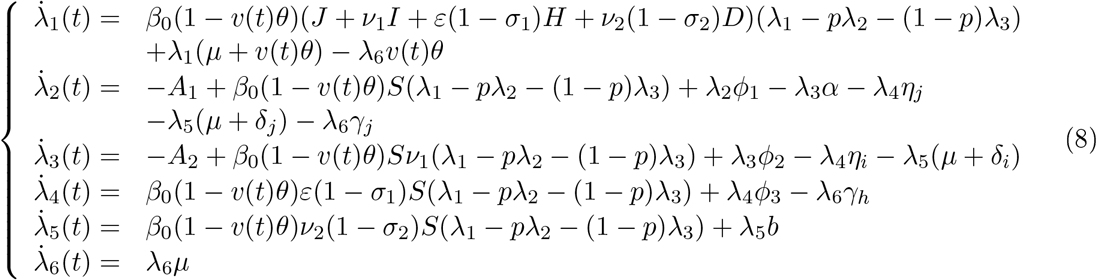

The optimal control and the associated state are found by solving the optimal problem, which consists of system (6) (with *υ*(*t*) = *υ*^*^, and non-negative initial condition), the adjoint system (8), the transversality conditions and the characterization of the control *υ^*^* given by:

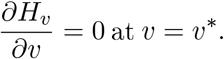

Thus, taking into account the bounds of *υ^*^* we get:

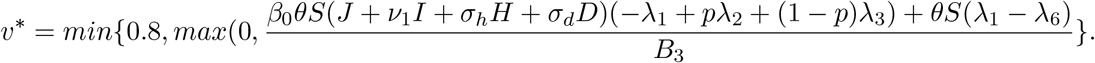

### 5.2 Combining optimal isolation and safe burial

The main objective of combining isolation and safe burial is to empower the reduction of EVD transmission by infected alive or dead individuals. We suppose in this paragraph that *σ*_1_ and *σ*_2_ are unkwon time dependent parameters. Thus, model (1) becomes:

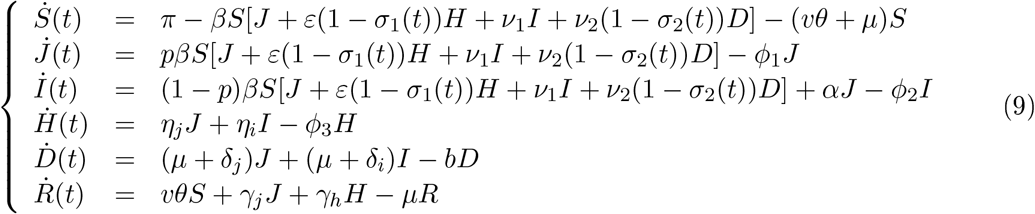

We define the cost functional that minimizes the alive infectious population by:

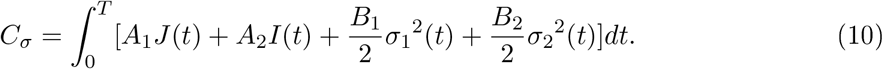

In (10), *T* is the maximum duration of implementation of isolation and safe burial, *A*_1_, *A*_2_ are the weights factor associated to infected with moderate and severe symptoms respectively, while *B*_1_, *B*_2_ are the weights factor linked to control variable *σ*_1_(*t*) and *σ*_2_(*t*), which are assumed Lebesgue measurable.

Let *x* be the state variable de_ned earlier in subsection 5.1. The optimal control problem here consists to find the optimal trajectory 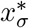 associated to the optimal control 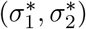 satisfying the control system (9) that minimizes the objective functional (10) over the set:

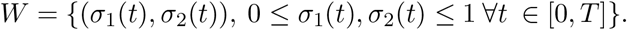

The proof of the existence and uniqueness of optimal control 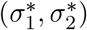 is similar to that of Theorem (5.1) and is skipped. However, the characterization of this control, given in the following theorem is determined in details in Appendix D from which we derive the main result as follows:

#### Theorem 5.2

*There exists* 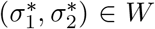 *so that the objective functional in* (10) *is minimized. Furthermore for small enough T*, 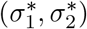 *is unique and is given by*

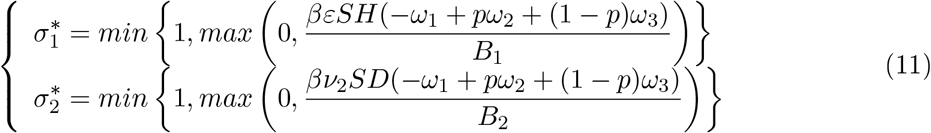

*where ω* = (*ω*_1_, *ω*_2_, *ω_3_, ω_4_, ω_5_, ω_6_), is a vector given in Appendix D*.

### 5.3 Numerical resolution of the optimal problems

In order to show the impact of optimal control measures, we solve the optimal problems defined in the subsections 5.1 and 5.2 by the forward-backward sweep method implemented in Matlab. Firstly, the state equations are simultaneously solved forward in time by a fourth-order Runge-Kutta method, with an initial guess for the control variables [31]. Secondly, the adjoint equations are also simultaneously solved backward in time using the solutions of the state equations. Finally, the controls are updated by inserting the new values of state and adjoint into its characterization and the process is repeated until the convergence occurs [14, 49].

We plot the curve of infected individuals with moderate symptoms, severe symptoms and hospitalized when the efforts are maximized during 50 days and consider the following two scenarios. First (i) constant vaccination is assumed: *υ* = 0.4 fli] (magenta curve in Figure 3), and (ii) optimal vaccination is considered (cyan curve in Figure 3). Secondly, (iii) the effectiveness of isolation and safe burial are assumed constant: *σ*_1_ = *σ*_2_ = 0.523 (red curve in Figure 4), and (iv) the effectiveness of isolation and safe burial are optimal (blue curve in Figure 4). These curves are plotted with the initial number of individuals *S, J, I, H, D* and *R* given in Table 3; and with the parameters values given in Table 2. The control profiles plotted in Figure 3 and 4 suggest that, the control function *υ*^*^ should be at the highest level during one week, before decreasing slowly to its lower bound at the final time, while 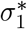, 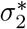 should be at the highest level during the first three days follow by a decrease. Precisely, after the first three days, the function *σ*_1_ decreases to 0.17, then increases to its upper bound, where it remains constant during about 20 days before dropping to its lower bound at the final time; whereas after three days, the control profile *σ*_2_ decreases to 0:2, then increases after to 0:61 and decreases again slowly to reach its lower bound. Despite the positive impact of these two optimal controls to reduce the disease burden, Figure 5 highlights that, optimal vaccination does better than the combination of isolation and safe burial. Therefore, one can recommend that, if for any disease outbreak, the vaccine is available, one should think of implementing large vaccination programs rather than any other control measures.

**Table 3:** Initial number of individuals for the optimal control implementation

| S(0) | J(0) | I(0) | H(0) | D(0) | R(0) |
| --- | --- | --- | --- | --- | --- |
| 6000 | 20 | 20 | 15 | 5 | 10 |

## 6 Conclusion and perspectives

In this paper, we proposed a deterministic model to understand the spread of Ebola virus disease which takes into account the fact that, the immune system of individuals differs from each other. The control reproduction number has been computed and its sensitivity analysis done and highlighted that, vaccination is the most important control measures at the beginning of the disease outbreak. A mathematical analysis of our model has been performed. Namely, we have shown the existence of a threshold value for the vaccination rate, which determines whether or not EVD dies out in population in the long run, by constructing suitable Lyapunov functions and using LaSalle’s Invariance Principle to prove that the disease-free and the endemic equilibria are globally asymptotically stable. Furthermore, we have first addressed optimal vaccination problem alone and secondly combined optimal isolation and safe burial problem, and show that, either strategy minimizes the number of infected individuals. The comparison of these control strategies has suggested that vaccination is the best. This might justify why, DRC authorities have adopted vaccination as the main control strategies during the 2018-2019 EVD outbreak in their country [77].

This work actually gives us the opportunity to explore other directions for the future:

- Investigate the optimal impacts of isolation, safe burial and vaccination all combined, to minimize the number of Ebola infected.
- Development of a similar model which include optimal contact tracing as another control strategy [9].
- Construction of an EVD model which incorporates control strategies and takes into account the transmission of EVD from animals to humans [8].
- Construction of a model taking into account the usefulness of media coverage and social networks.

## Data Availability

N/A

## Acknowledgments

The second author(BT), acknowledges the financial support of the University of Pretoria Senior Postdoctoral Program Grant (2018-2020).

## Appendix A: Proof of Theorem 2.2

This proposition is shown using the theory of monotone systems and the comparison principle. We begin by proving that, if *S*(0) > 0 then ∀ *t* ≥ 0, *S*(*t*) > 0.

**Figure 3:**
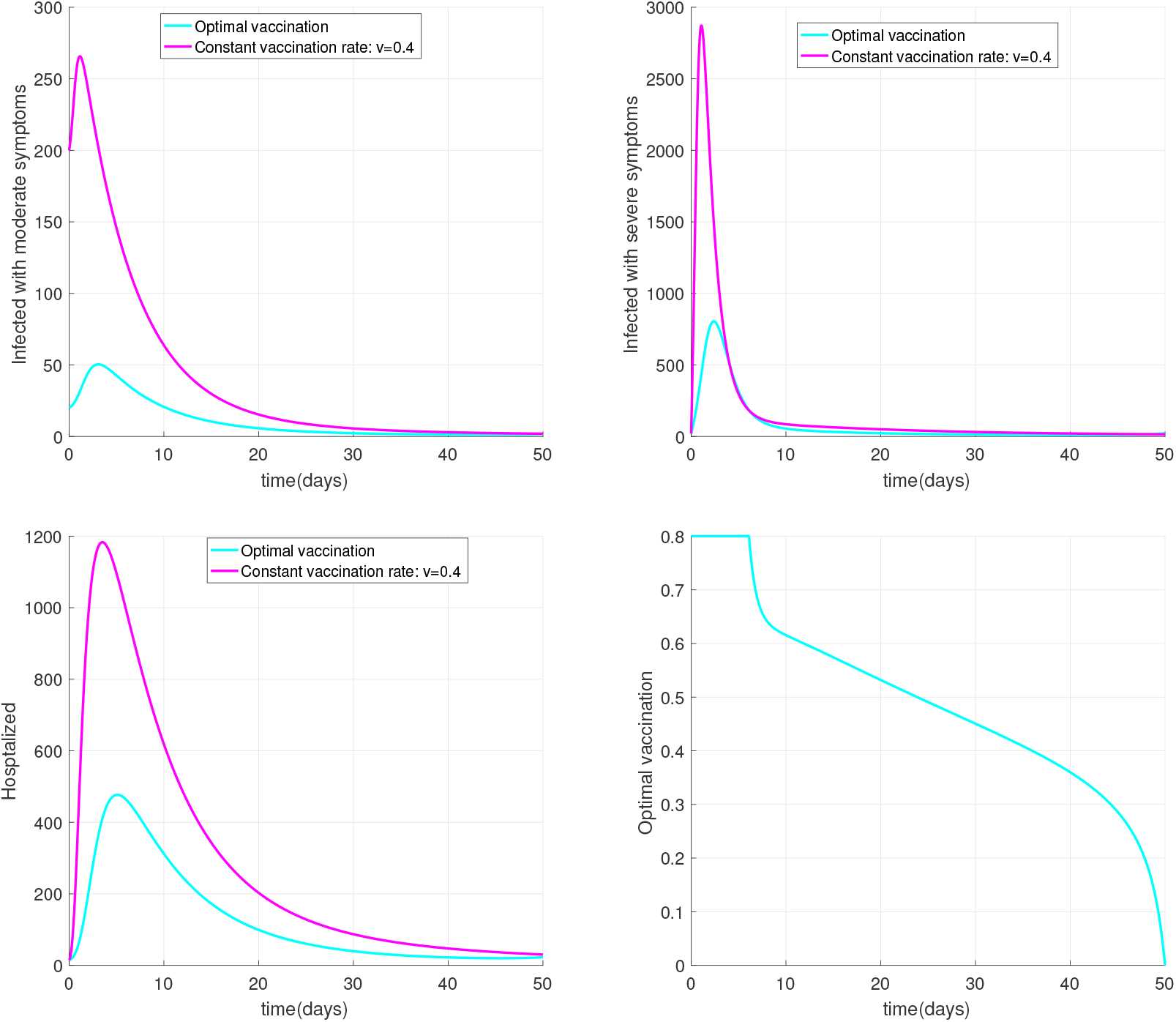
This Figure gives the curves for the time evolution of infected individuals with moderate symptoms, severe symptoms and hospitalized when (i) the vaccination is constant: *υ* = 0.4 [12] (magenta curve), (ii) the vaccination is optimal (cyan curve), with the parameters: *A*_1_ = 0.02, *A*_2_ = 0.015, *B*_3_ = 1 (the other parameters are as in Table 2).

Let’s suppose *S*(0) > 0, then from the first equation of (1), if

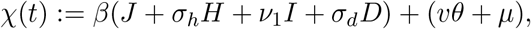

the integration from 0 to *t* > 0 gives

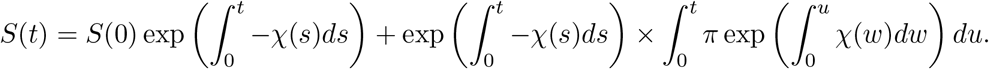

Therefore *S*(*t*) > 0, ∀ *t* ≥ 0.

To prove that *J*(*t*) ≥ 0,*I*(*t*) ≥ 0,*H*(*t*) ≥ 0,*D*(*t*) ≥ 0,*R*(*t*) ≥ 0 ∀ *t* ≥ 0, when *J*(0) ≥ 0,*I*(0) ≥ 0,*H*(0) ≥ 0,*D*(0) ≥ 0 and *R*(0) ≥ 0.

Let’s defne the function *R*_1_ such as

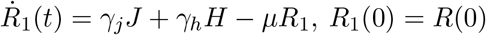

**Figure 4:**
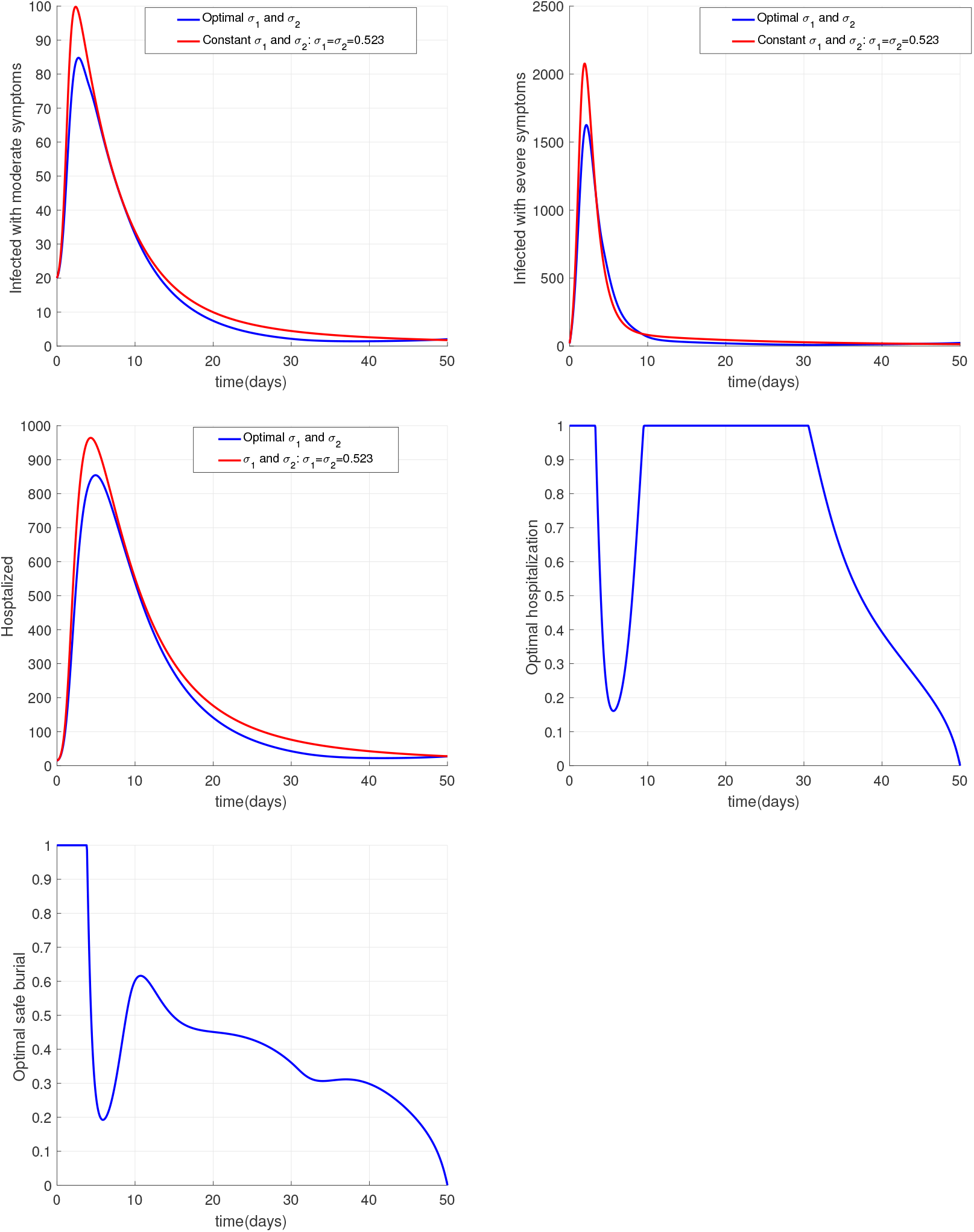
This Figure plots the curves for the time evolution of infected individuals with moderate symptoms, severe symptoms and hospitalized when: (i) the efectiveness of isolation and safe burial are constant: *σ*_1_ = *σ*_2_ =0.523 (red curve); (ii) the efectiveness of isolation and safe burial are optimal (blue curve), with the parameters *A*_1_ =0.02,*A*_2_ =0.015,*B*_1_ = *B*_2_ =1 (the other parameters are as in Table 2).

**Figure 5:**
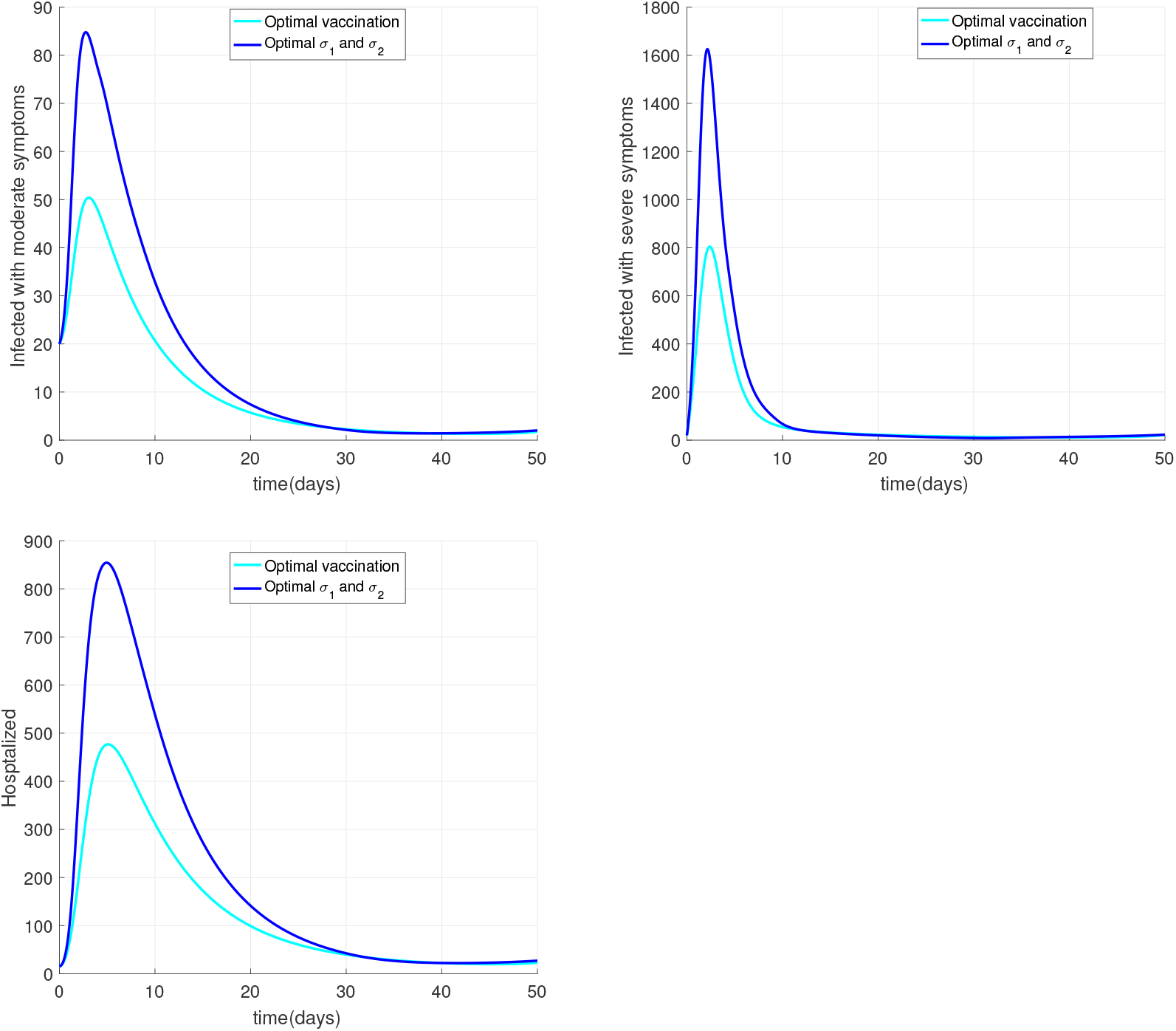
This Figure compares the curves of the time evolution infected individuals with moderate symptoms, severe symptoms and hospitalized when: (i) the effectiveness of isolation and safe burial are optimal (blue curve); (ii) the vaccination is optimal (cyan curve) with the parameters of the figures above.

and consider the following sub-equations related to the variables *J, I, H,D* and *R*_1_.

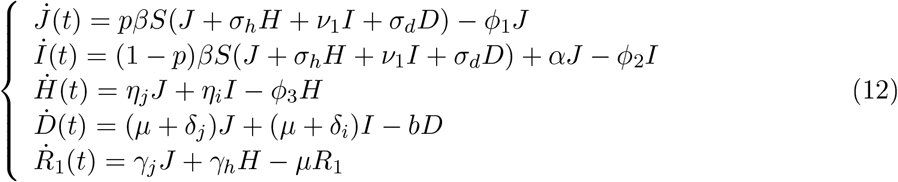

System (12) takes the matrix form:

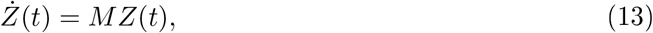

where,

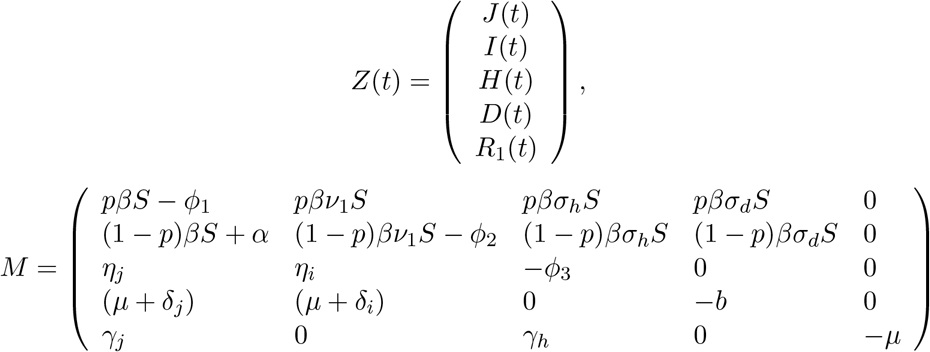

It is straightforward that *M* is a Metzler matrix. Thus, (13) is a monotone system. It follows that, 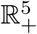is invariant under the flow of (13). Therefore, *J*(*t*) ≥ 0,*I*(*t*) ≥ 0,*H*(*t*) ≥ 0,*D*(*t*) ≥ 0, and *R*_1_(*t*) ≥ 0, ∀ *t* ≥ 0.

Moreover, the application of the comparison theorem [60, 64] leads to *R*(*t*) ≥ *R*_1_(*t*) ∀ *t* ≥ 0. Hence, *R*(*t*) ≥ 0 ∀ *t* ≥ 0, and the proof is achieved.

## Appendix B: Proof of Theorem 2.6

Let (*S*^*^,*J*^*^,*I*^*^,*H*^*^,*D*^*^,*R*^*^) be an equilibrium point of (3). Setting the right hand side of (3) to zero we obtain.

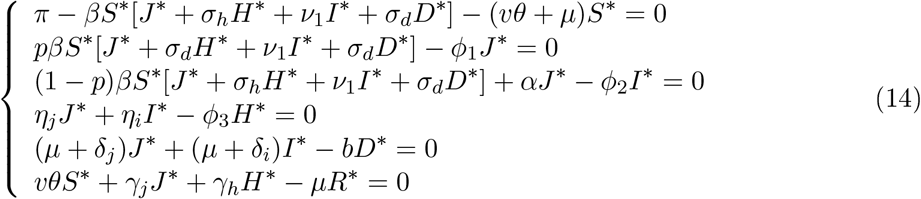

For simplifcation, set

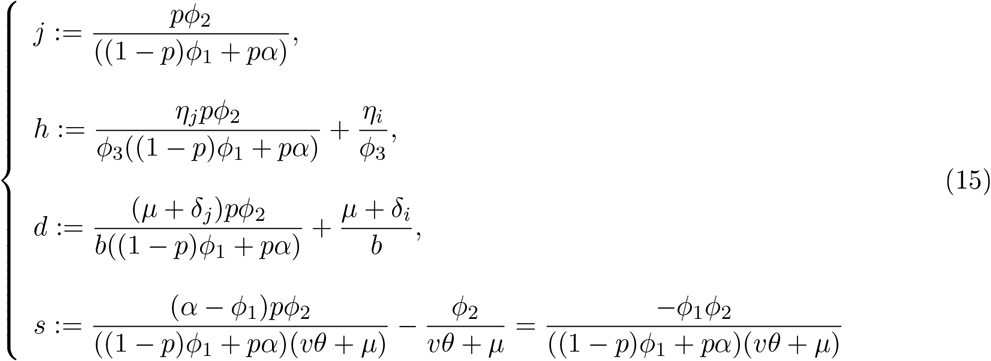

From the second and the third equations of (14), we get:

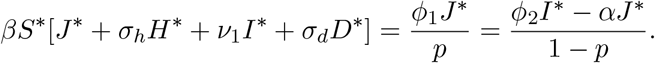

Therefore, from the fourth and the fifth equation equation of (14), it is straightforward that

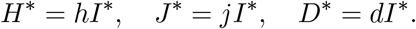

The sum of the first three equations of system (14) gives

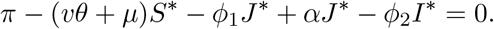

Hence,

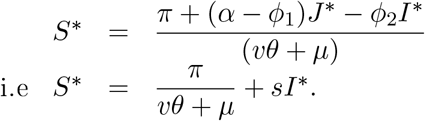

Plugging the latter expressions of *J*^*^,*I*^*^ and *D*^*^ into the first equation of (14) gives

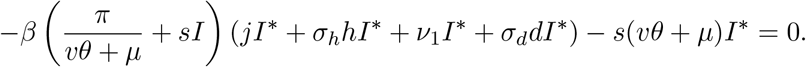

Thus, *I*^*^ =0 (corresponding to the disease-free equilibrium point) or

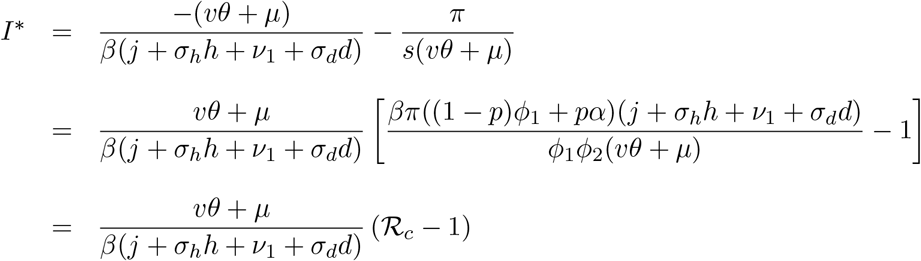

where,

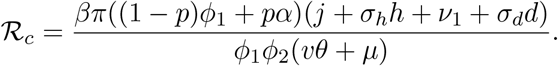

Thus, if 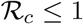, model (1) has a unique equilibrium, namely the disease-free equilibrium (DFE). Conversely, if 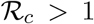 it has two equilibria: the DFE *E*0 and a unique endemic equilibrium *E*_1_ =(*S*^*^,*J*^*^,*I*^*^,*H*^*^,*D*^*^,*R*^*^) where

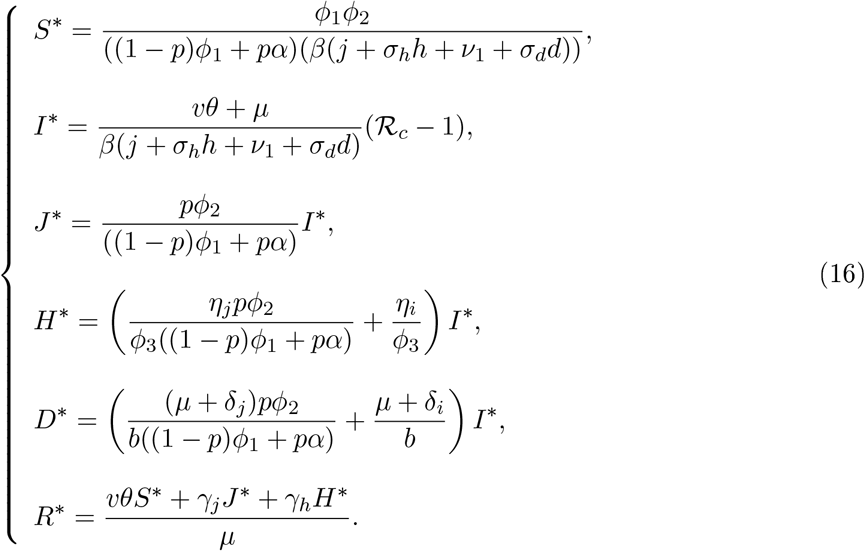

## Appendix C: Proof of the global stability of equilibria

### Proof of Theorem..1

Since the variable *R* does not appear in the remaining equation of (3), its equation is decoupled, thus reducing the dynamics of (3) to that of the following system

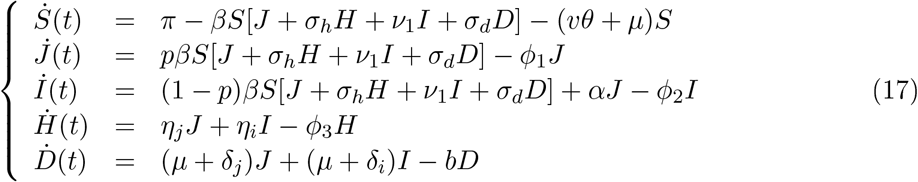

Let’s recall that 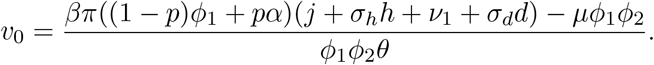.

It is straightforward that,

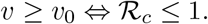

Thus, to prove the global asymptotic stability of the DFE for model (3) when *υ* ≥ *υ*_0_, it sufces to prove the global asymptotic stability of the DFE for model (17) when 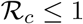.

To construct a Lyapunov function for the DFE of (17) when 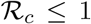, we use the matrix-theoretic method as described in [63].

Let’s consider the infectious compartments *J, I, H, D* and the non infectious compartment *S*. Set *x*:= (*J, I, H, D*) and *y*:= *S*. Following [70], we define

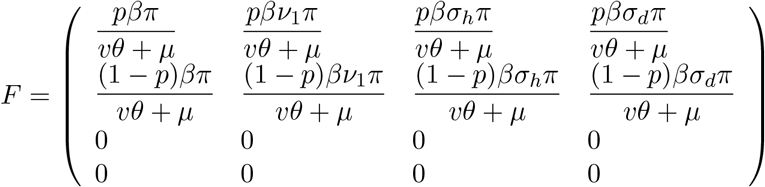

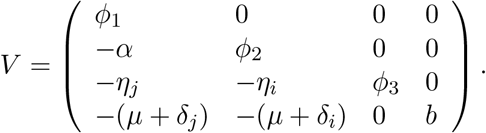

So,

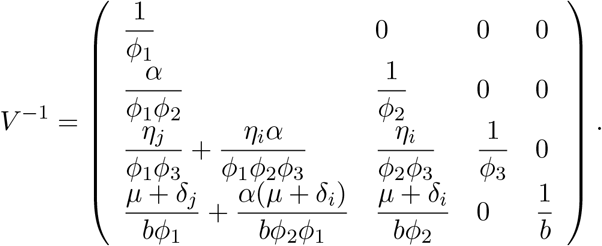

Thus, the control reproduction number 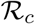 for model (3) is given by 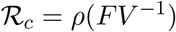 [70]. Following [63], set

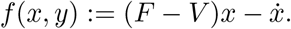

Then,

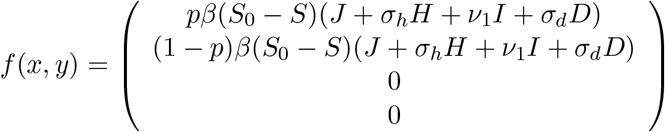

where 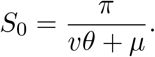

It is straightforward that *f*(*x, y*) ≥ 0 in Ω.

Simple calculations give

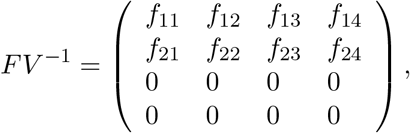

where

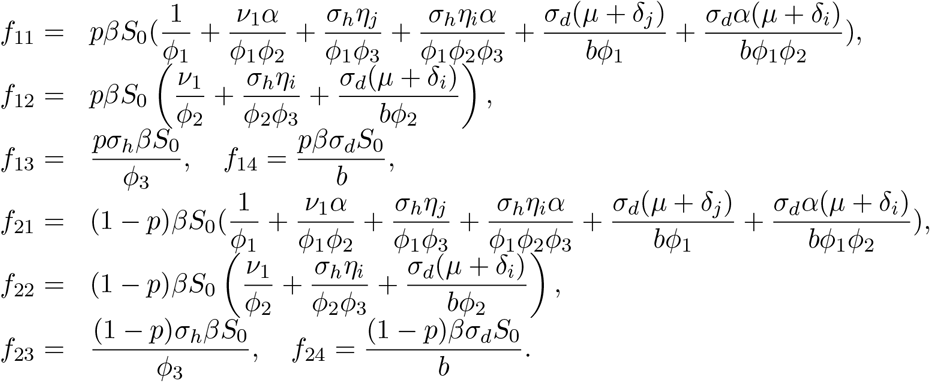

A left eigenvector *w* =(*w*_1_,*w*_2_,*w*_3_,*w*_4_)*^T^* of the eigenvalue 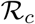 of the next generation matrix FV ^−1^ is given by:

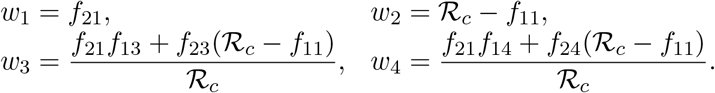

One can easily show that 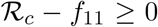, then following [63], the function *L* = *w^T^V*^−1^*x* is a Lyapunov function for the DFE of model (3) defned in Ω.

In fact, differentiating *L* along the solutions of model (3) gives:

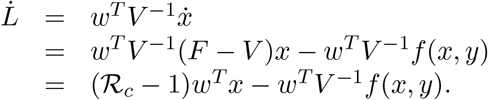

Since *w^T^* > 0, *V*^−1^ > 0 and *f*(*x, y*) ≥ 0 in Ω, we have 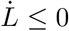 when 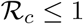.

Moreover,

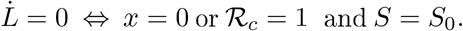

Thus, the largest invariant subset contained in the set 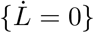 is 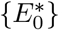, where 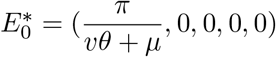.

Therefore, from LaSalle Invariance Principle [57]

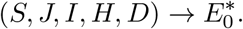

Hence, the DFE of model (17) is GAS, and so, *E*_0_ is globally asymptotically stable.

### Proof of Theorem 4.2

The expression of 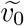 is given by

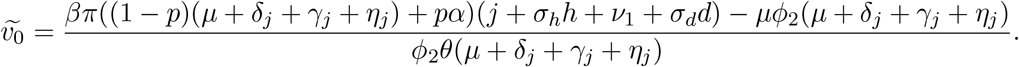

Let’s remark that, 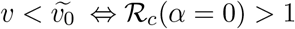.

Thus, it suffices to prove the global asymptotic stability of the endemic equilibrium for model (17), when 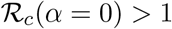. Our proof here is similar to the ones proposed in [8, 63].

Set

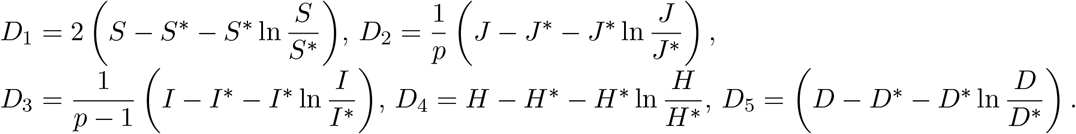

Then,

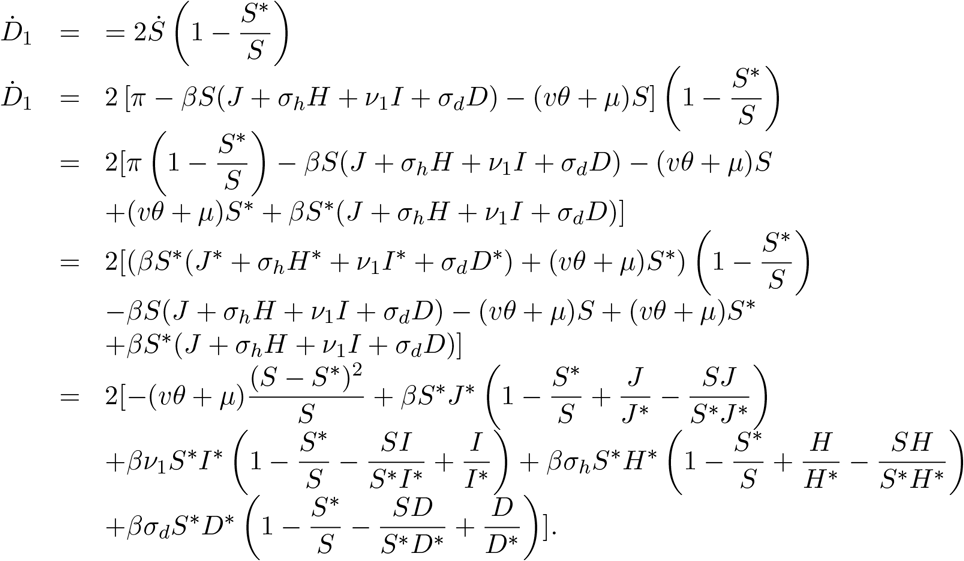

Note that 1 − *x* + ln *x* ≤ 0 for *x*> 0. Hence,

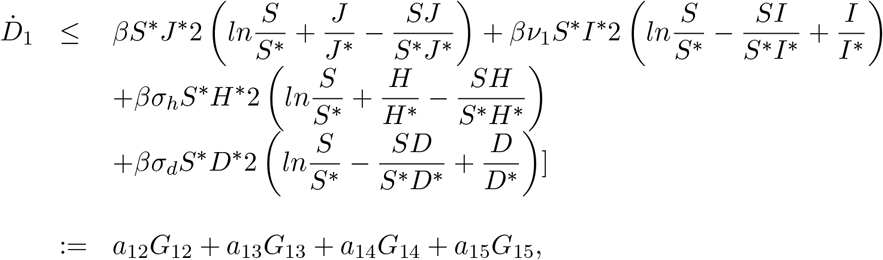

with,

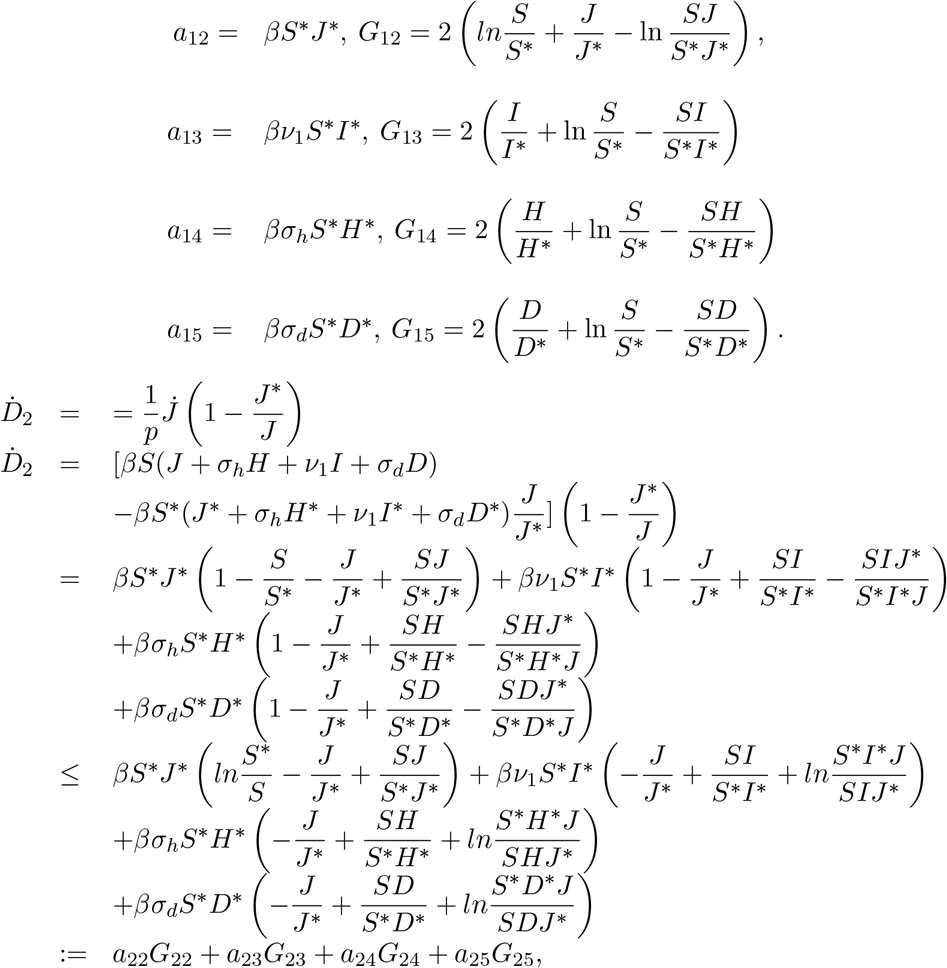

where,

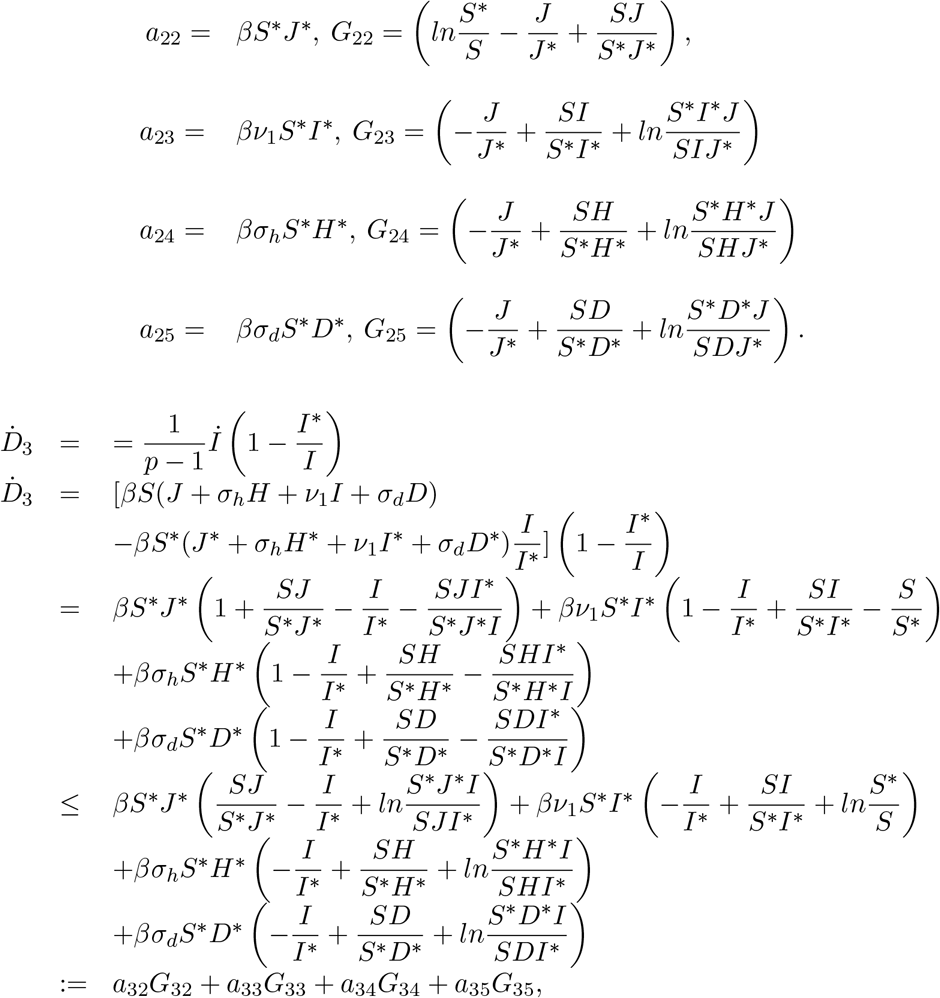

and,

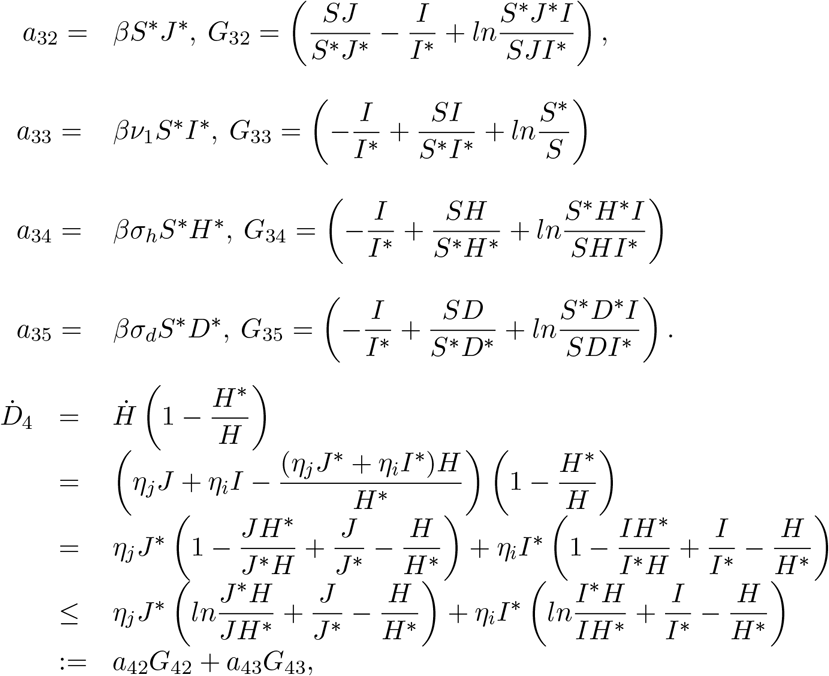

with,

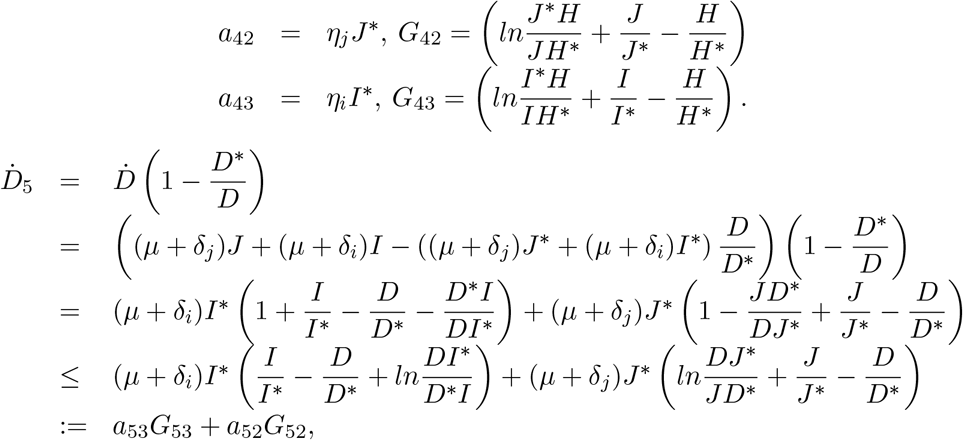

and

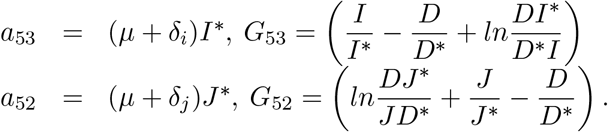

Simple computations yield

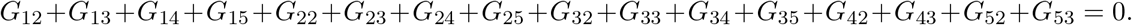

Thus, there exists (*c_i_*)_1≤_*_i_*_≤5_,*c_i_* ≠ 0, such that 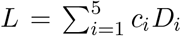 is a Lyapunov function [63]. Moreover, it is straightforward that, the largest invariant subset such that 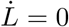 is the singleton *E*_1_. Thus, by LaSalle Invariance Principle, *E*_1_ is GAS.

## Appendix D: Determination of the characterization of 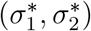

Following the Pontryagin’s maximum principle, there exists a function

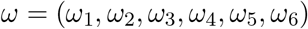

*ω* =(*ω*_1_,*ω*_2_,*ω*_3_,*ω*_4_,*ω*_5_,*ω*_6_)
called the adjoint vector so as

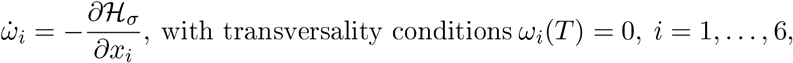

where 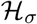 is the Hamiltonian defined by:

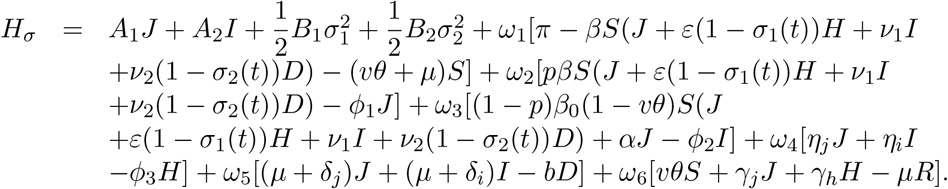

The adjoint variables satisfy the system:

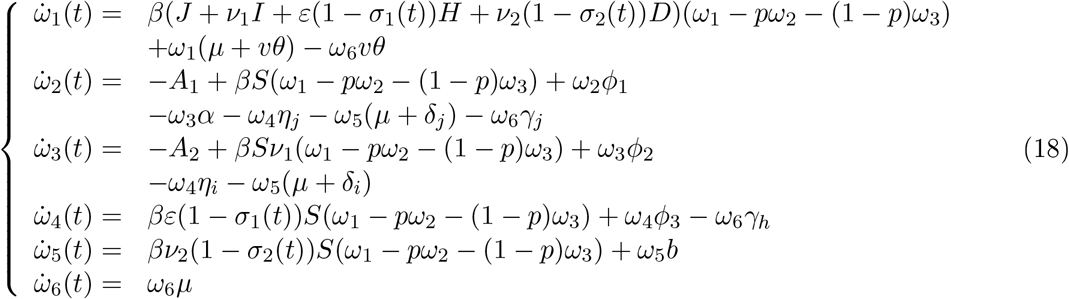

The optimal control and the associated state are found by solving the optimal problems, which consists of system (9) (with 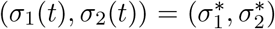, and non-negative initial condition), the adjoint system (18), the transversality conditions and the characterization of the control 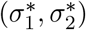 given by [49]:

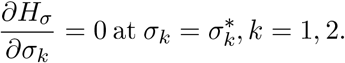

Thus, 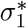 and 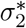 are given by:

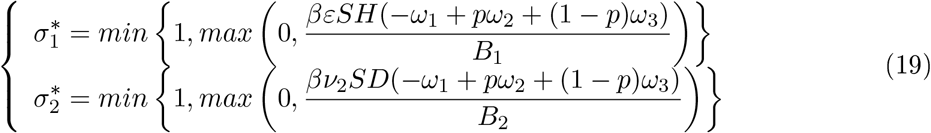

## References

[1] F. B. Agusto, Mathematical model of Ebola transmission dynamics with relapse and reinfection, Math. Biosci., 283 (2017), 48–59.

[2] F. B. Agusto, M. I. Teboh-Ewungkem, A. B. Gumel, Mathematical Assessment of the Effect of Traditionnal Beliefs and Customs on the Transmission Dynamics of the 2014 Ebola outbreaks, BMC Medecine, 13:96 (2015), 1–17.

[3] M.D. Ahmad, M. Usman,A. Khan, M. Imran, Optimal Control Analysis of Ebola disease with Control Strategies of Quarantine and Vaccination, Infectious Diseases of Poverty, 5:72 (2016), 1–12.doi:10.1186/s40249-016-0161-6.

[4] I. Area, J. Losada, F. Ndaïrou, J. J. Nieto, D. D. Tcheutia, Mathematical Modeling of 2014 Ebola Outbreak, Math. Meth. Appl. Sci. 40 (2015) 6114–6122.

[5] I. Area, F. Ndaïrou, J. J. Nieto, C. J. Silva, D. F. M. Torres, Ebola Model and Optimal Control with Vaccination Constraints, JIMO, 278 (2017), 1–21.

[6] T. Berge, S. Bowong, J. M.-S. Lubuma, M. L. M. Manyombe, Modelling Ebola Virus Disease Transmissions with Reservoir in a Complex Virus Life Ecology, Math. Biosci., 15 (1) (2018), 21-56.

[7] T. Berge, M. Chapwanya, J. S.-M. Lubuma, Y. A. Terefe, A Mathematical Model for Ebola Epidemic with Self-Protection Measures, J. Biol. Syst., 11 (1):(2016), 42-74.

[8] T. Berge, J. S.-M. Lubuma, A. J. O. Tassé, H. M. Tenkam, Dynamics of Host-Reservoir Transmission of Ebola with Spillover Potential to Humans, EJQTDE, 14(2018), 1-32. https://doi.org/10.14232/ejqtde.2018.1.1.4.

[9] T. Berge, A. J. O. Tassé, H. M. Tenkam, Mathematical modeling of contact tracing as a control strategy of Ebola Virus Disease, IJB, 11 (2018), 1850093–1-36.

[10] Berman, Plemmons, Nonnegative Matrices in the Mathematical Sciences Academic, Press, New York (1979).

[11] J. C. Blackwood, L. M. Childs, The Role of Interconnectivity in Control of an Epidemic, Scientific Reports, 6:29263 (2016), 1–10.

[12] E. N. Bodine, C. Cook, M. Shorten, The Potential impact of a Prophylactic Vaccine for Ebola virus disease in Sierra Leone, Math. Biosci, 15 (2018), 337–359.

[13] C. S. Bornaa, O. D. Makinde, I. Y. Seini, Eco-Epidemiological Model and Optimal Control of Disease Transmission Between Humans and Animals, Commun. Math. Biol. Neurosci. 2015:26 (2015) 2052–2541.

[14] B. Buonomo, D. Lacitignola, C. Varga-De-Leön, Qualitative Analysis and Optimal Control of an Epidemic Model with Vacciation and Treatment, Math Comput Simulat, 100 (2014), 88–102.

[15] D. Branigan, Evidence shows Ring Vaccination Strategy Effective in Limiting Ebola Outbreak in DRC, Health Policy Watch, 15-04-2019, https://www.healthpolicy-watch.org/evidence-shows-ring-vaccination-strategy-effective-in-limiting-ebola-outbreak-in-drc

[16] D. Butler, How Disease Detectives are fighting Ebola’s Spread, nature, www.nature.com/news/how-disease-detectives-are-fighting-ebola-s-spread-1.16069, 02 October 2014.

[17] G. Caleo, J. Ducombe, F. Jephcott, K. Lokuge, C. Mills, E. Looijen, F. Theorharaki, R. Kremer, K. Kleijer, J. Squire, M. Lamin, B. Stringer, H. A. Weiss, D. Culli, G. L. D. Tama, J. Greig, The Factors Affecting the Household Transmission Dvamics and Community Compliance with Eola Control Measures: a Mixed-Method Study in a Rural Village in Sierra Leone, BMC Public Health, 18:248 (2018), 1–13.

[18] A. Camacho, R. M. Eggo, S. Funk, C. H. Watson, A. J. Kucharski, Estimating the Probability of Demonstrating Vaccine Efficacy in the Declining Ebola Epidemic: A Bayesian Modelling Approach, BMj Open, 5:e009346 (2015), 1–6.

[19] A. Camacho, A. J. Kucharski, S. Funk, J. Breman, P. Piot, W. J. Edmunds, Potential for Large Outbreaks of Ebola Virus Disease, Epidemics, 9 (2014), 70–78.

[20] S. J. Chapman, V. S. Hill, Human Genetic Susceptibility to Infectious Disease, Nature Reviews Genetics, 13(3) (2012), 175-188.

[21] J. R. Conrad, L. Xue, J. Dewar, J. M. Hyman, Modeling the impact of Behavior Change on the Spread of Ebola, Mathematical and Statistical Modeling for Emerging and Re-emerging Infectious Diseases, doi: 10.1007/978-3-319-40413-4_2, (2016).

[22] Crisis update - April 2020,MSF, 23 April 2020, https://www.msf.org/drc-ebola-outbreak-crisis-update.

[23] A. Dénes, A. B. Gumel, Modeling the Impact of Quarantine During an Outbreak of Ebola Virus Disease, Infectious Disease Modelling, 4 (2019) 12-27.

[24] T. S. Do, Y. S. Lee, Modeling the Spread of Ebola, Osong Public Health Res Perspect 7(1) (2016), 43-48.

[25] J. M. Drake, R. B. Kaul, L. W. Alexander, S. M. O. Regan, A. M. Kramer, J. T. Pulliam, M. J. Ferrari, A. W. Park, Ebola Cases and Health System Demand in Liberia, Plos Biology, 13(1) (2015), 1-20, doi: 10.1371/journal.pbio.1002056.

[26] M. O. Durojaye, I. J. Ajie, Mathematical Model of the Spread and Control of Ebola Virus Disease, J. Appl. Math. Com,put, 7 (2017), 23–31.

[27] L. Garrett, Ebola: Story of an Outbreak, Hachette Books, New York (2014).

[28] D. Gao, Transmission Dynamics of some Epidemiological Patch Model, University of Miami, (2012), phd thesis.

[29] A. Gideon, I. Miranda, Teboh-Ewungkem, A Mathematical Model with Quarantine states for the dynamics of Ebola Virus Disease in Human Populations, Comput. Math. Methods Med., 2016 (2016), 1–29.

[30] J. R. Glynn, H. Bower, S. Johnson, C. F. Houlihan, C. Montesano, J. T. Scott, M. G. Semple, M. Bangura, A. J. Kamara, O. Kamara, S. H. Mansaray, D. Sesay, C. Turay, S. Dicks, R. E. G. Wadoum, V. Colizzi, F. Checchi, D. Samuel, R. S. Tedder, Asymptomatic Infection and Unrecognised Ebola Virus Disease in Ebola-Affected Households in Sierra Leone: A Cross-Sectional Study Using a New Non-Invasive Assay for Antibodies to Ebola Virus, Lancet Infect Dis, 17 (2017), 645–53.

[31] J. N. C. Goncalves, Epidemiological Models and Optimal Control Theory-Applications to Marketing and Computer Viruses Transmission, Universidade do Minho, (2017), phd thesis.

[32] E. Grigorieva, E. Khailov, Determination of the Optimal Controls for an Ebola Epidemic Model, DCDS-S, 11 (2018), 1071–1101.

[33] Hal Smith, An Introduction to Delay Differential Equations with Applications to the Life Sciences, Springer, 2010, p.154-155.

[34] J. M. Hyman, J. Lie, E. A. Stanley, Modeling the Impact of Random Screening and Contact Tracing in Reducing the Spread of HIV, Mathematical Biosciences, 181 (2003), 17–54.

[35] M. Imran, A. Khan, A. R. Ansari, Modeling Transmission Dynamics of Ebola Virus Disease, IJB, 10 (2017), 1750057–1–35.

[36] K. Kabli, S. E. Moujaddid, K. Niri, A. Tridane, Comparative System Analysis of the Ebola Virus Epidemic Model, Infectious Disease Modelling, 3 (2018), 145–159.

[37] S. Kim, A. Tridane, D.E. Chang, Human Migration and Mosquito-Borne Diseases in Africa, Math Popul Stud, 23 (2016), 123–146.

[38] M. A. Kimaro, E. S. Massawe, D. O. Makinde, Modelling the Optimal Control of Transmission Dynamics of Mycobacterium Ulceran Infection, OJEpi 5(2015) 229–243.

[39] A. J. Kucharski, R. M. Eggo, C. H. Watson, A. Camacho, S. Funk, J. Edmungs, Effectiveness of Ring Vaccination as Control Strategy for Ebola Virus Disease, Emerg. Inf. Dis., 22 (2016), 105–108.

[40] J. Legrand, R. F. Grais, P. Y. Boelle, A. J. Valleron, A. Flahault, Understanding the Dynamics of Ebola Epidemics, Epidemiol. Infect., 135 (2007), 610–621.

[41] S. Lenhart, J.T. Workman, Optimal Control Applied to Biological Models, Mathematical and Computational Biology Series, Chapman & Hall/CRM, (2007).

[42] E. M. Leroy et al., Human asymptomatic ebola infection and strong inflammatory response, Lancet 355 (2000), 2210–2215.

[43] Li Li, Transmission Dynamics of Ebola Virus Disease with Human Mobility in Sierra Leone, Chaos, Solitons and Fractals, 104 (2017), 575–579.

[44] Y. Li, H. Wang, Y-G. Wang, Experiences and Challenges in the Health Protection of Medical Teams in the Chinese Ebola Treatment Center, Liberia: a Qualitative Study, Infect Dis Poverty, 7:92 (2018), 1–12.

[45] C. E. Madubueze, A. R. Kimbir, T. Aboiyar, Global Model with Contact Tracing and Quarantine, AAM 13 (2018), 382–403.

[46] D. F. Maron, Why does Ebola Keep Showing Up in the Democratic Republic of the Congo?, Scientific American, (2018), https://www.scientificamerican.com/article/why-does-ebola-keep-showing-up-in-the-democratic-republic-of-the-congo/.

[47] S. Merler, M. Ajelli, L. Fumanelli, S. Parlamento, A. P. Y. Piontti, N. E. Dean, G. Putoto, D. Carraro, I. M. Longini, J. M. E. Halloran, A. Vespignani, Containing Ebola at the Source with Ring Vaccination, PLOS Negl. Trop. Dis, 10 (11), (2016), 1-11.

[48] S. D. D. Njankou, Mathematical Models of Ebola Virus Disease with Socio-Economics Dynamics, University of Stellenbosch, South Africa, (2019), phd thesis.

[49] O. O. Onyejekwe, E. Z. Kebede, Epidemiological Modeling of Measles Infection with Optimal Control of Vaccination and Supportive Treatment, ACM, 4(4) (2015).

[50] Oxford Vaccine Group, Vaccine Knowledge Project, University of Oxford, 29-06-2018, vk.org.ox.ac.uk./vk/disease-vaccinated-populations.

[51] J. Ponce, Y. Zheng, G. Lin, Z. Feng, Assessing the Effects of Modeling the Spectrum of Clinical Symptoms on the Dynamics and Control of Ebola, J. Theor. Biol 467 (2019) 111–122.

[52] J. Prescott et al, Postmorten Stability of Ebola Virus, Emerg. Inf. Dis., 21 (2015).

[53] A. Rachah, D. F. M. Torres, Analysis, Simulation and Optimal Control of a SEIR Model for Ebola Virus with Demographic Effects, DDNS, (2017),

[54] A. Rachah, D. F. M. Torres, Optimal Control Strategies for the Spread of Ebola in West Africa, J. Math. Anal, 7 (2016), 102–114.

[55] A. L. Rasmussen, A. Okumara, M. T. Ferris, R. Green, F. Feldmann, S. M. Kelly, D. P. Scott, D. Safronetz, E. Haddock, R. LaCasse, M. J. Thomas, P. Sova, V. S. Carter, J. M. Weiss, D. R. Miller, G. D. Shaw, M. J. Korth, M. T. Heise, R. S. Baric, F. Villena, M. G. Katze, Host Genetic Diversity Enables Ebola Haemorrhagic Fever Pathogenesis and Resistance, Science, 346(6212) (2014), 987-991.

[56] C. M. Rivers, E. T. Lofgren, M. Marathe, S. Eubank, B. L. Lewis, Modelling the Impact of Interventions on an Epidemic of Ebola in Sierra Leone and Liberia, PLOS currents Outbreaks, DOI: 10.1371/currents.Outbreaks.fd38dd85078565450b0be3fcd78f5ccf, (2014), 1-19.

[57] M. A. Safi, Mathematical Analysis of the Role of Quarantine and Isolation in Epidemiology, University of Manitoba, (2010), phd thesis.

[58] M. A. Safi, A. B. Gumel, Dynamics Analysis of a Quarantine Model in two Patches, Math.Meth.Appl.Sci, 38 (2015), 349–364.

[59] D. Salem, R. Smith, A Mathematical Model of Ebola Virus Disease: Using Sensitivity Analysis to Determine Effective Intervention Targets, SCS, (2016), 16-23.

[60] M. Salmani. P.van den Driessche, J. Watmough, A Model for Disease Transmission in a Patchy Environment, Disc, and Cont. Dyna. Syst., 6 (2006), 185–202.

[61] A. Sanchez, K. E. Wagoner, P. E. Rollin, Sequence-Based Human Leukocyte Antigen-B Typing of Patients Infected with Ebola Virus in Uganda in 2000: Identification of Alleles Associated with Fatal and non Fatal Disease Outcomes, The Journal of Infectious Diseases, 196 (2007), S329-336.

[62] M. Shen, Y. Xiao, L. Rong, Modeling the Effect of Comprehensive Interventions of Ebola Virus Transmission, srep, 5.15818(2015), 1-14.

[63] Z. Shuai, P. Van den Driessche, Global Stability of Infections Disease Models Using Lyapunov Functions; J. Appl. Math, 73 (2013), 1513–1532, http://www.siam.org/journals/siap/73-4/87664.html.

[64] H. L. Smith, P. Waltman, The Theory of the Chemostat, Cambridge University Press, New York, 1995.

[65] H. R. Thieme, Persistence under Relaxed Point-Dissipativity (with Applications to an Endemic Model), J. Math. Anal. 24 (1993) 407–435.

[66] T. W. Tulu, B. Tian, Z. Wu, Modeling the Effect of Quarantine and Vaccination on Ebola Disease, Adv. Differ. Equ 2017:178 (2017) 1–14.

[67] S. Towers, O. Patterson-Lomba, C. Castillo-Chavez, Temporal Variations in the Effective Reproduction Number of the 2014 West Afica Ebola outbreak, PLOS Currents Outbreaks, doi:10.1371/currents.outbreaks.9e4c4294ec8celadad283172bl6bc908, (2014), 1-13.

[68] B. Tsanou, S. Bowong, J. Mbang, J. Lubuma, Assessing the Impact of the Environmental Contamination on the Transmission of Ebola Virus Disease (EVD). J. Appl. Math. Comput., 55 (2017), 205–243.

[69] B. Tsanou, J. M.-S. Lubuma, G. M. Moremedi, N. Morris, R. Kondera-Shava, A Simple Mathematical Model for Ebola in Africa; J Biol Dyn, 11:1 (2016), 42–74.

[70] P. Van den Driessche, J. Watmough, Reproduction Numbers and Sub-threshold Endemic Equilibria for Compartment Models of Disease Transmission, Math. Biosci, 180 (2002), 29–48.

[71] J. Wang, C. Modnak, Modeling Cholera Dynamics with Controls, CAMQ, 19 (2011), 255–273.

[72] X.-S. Wang and L. Zhong, Ebola outbreak in West Afica: Real-time estimation and multiple-wave prediction, Math. Biosci, 12 (5) (2015), 1055-1063.

[73] WHO, Ebola Virus Disease, (2019), https://www.who.int/news-room/fact-sheets/detail/ebola-virus-disease.

[74] WHO, Ebola Virus Disease-Democratic Republic of Congo, 08-08-2019, https://www.who.int/crs/don/08-august-2019-ebola-drc/en/.

[75] WHO, Emergencies Preparedness, Response: Factors that Contributed to Undetected Spread of the Ebola Virus and Impeded Rapid Containment, (2015), https://www.who.int/csr/disease/ebola/one-vear-report/factors/en.

[76] WHO, Global Vaccine Safety, https://www.who.int/vaccine_safety/initiative/detection/immunization_misconception/

[77] WHO, Public health round-up, Bull World Health, 97(12) (2019), 793–794.

[78] WHO, Second Ebola Vaccine to Complement “Ring Vaccination” given Green Light in DRC, 23-09-2019, https://www.who.int/news-room/detail/23-09-2019-second-ebola-vaccine-to-complement-ring-vaccination-given-green-light-in-drc.

[79] WHO, Vaccinations against Ebola begin in DR Congo City of Goma, AlJAZEERA, 22-07-2019, https://www.google.com/amp/s/www.aljazeera.com/amp/news/2019/07/vaccinations-ebola-dr-congo-city-goma-19071511221895.html.

[80] WHO, WHO adapts Ebola Vaccination Strategy in the Democratic Republic of Congo to account for Insecurity and Community Feedback, 07-05-2019, https://www.who.int/news-room/detail/07-05-2019-who-adapts-ebola-vaccination-strategv-in-the-democratic-republic-of-congo-to-account-for-insecurity-and-communitv-feedback.

[81] S. Wiafe, E. Veilleux-Gravel, R. Smith, Using Sensitivity Analysis to Examine the Effects of an Ebola Vaccine, SCS, 12 (2017), 61–72.

[82] F. K. Zadeh, A. V. Griensven, J. Nossent, Parameter Screening: The Use of a Dummy Parameter to Identify Non-Infiuential Parameter in a Global Sensitivity Analysis, Geophysical Research Abstracts, 19 (2017), https://www.researchgate.net/publication/316548214.

[83] J-M. Zhu, L. Wang, J-B. Liu, Eradication of Ebola Based on Dynamic Programming, Com,put. Math. Methods Med., 2016 (2016), 1–9.

